# Effects of underlying morbidities on the occurrence of deaths in COVID-19 patients: A systematic review and meta-analysis

**DOI:** 10.1101/2020.05.08.20095968

**Authors:** Md. Mostaured Ali Khan, Md. Nuruzzaman Khan, Md. Golam Mustagir, Juwel Rana, Md. Saiful Islam, Md. Iqbal Kabir

## Abstract

**Background:** Coronavirus disease 2019 (COVID-19), the most hectic pandemic of the era, is increasing exponentially and taking thousands of lives worldwide. This study aimed to assess the prevalence of pre-existing morbidities among COVID-19 infected patients and their mortality risks against each type of pre-existing morbidity category.

**Methods:** To conduct this systematic review and meta-analysis, Medline, Web of Science, Scopus, and CINAHL databases were searched using specified relavent keywords. Further searches were conducted using the reference list of the selected studies, renowned pre-print servers (e.g., medRxiv, bioRixv, SSRN), and relevant journal websites. Studies written in the English language included if those were conducted among COVID-19 patients with and without comorbidities and presented survivor vs. non-survivor counts or hazard/odds of deaths or survivors against types of pre-existing morbidities. Comorbidities reported in the selected studies were grouped into eight categories. The pooled likelihoods of deaths in each category were estimated using a fixed or random-effect model, based on the heterogeneity assessment. Publication bias was assessed by visual inspection of the funnel plot asymmetry and Egger’s regression test. Trim and Fill method was used if there any publication bias was found.

**Results:** A total of 42 studies included in this study comprised of 39,398 samples. The most common pre-existing morbidities in COVID-19 infected patients were hypertension (36.5%), cardiovascular disease (11.9%), and diabetes (22.0%). The higher likelihood of deaths was found among COVID-19 patients who had pre-existing cardiovascular diseases (OR: 3.32, 95% CI: 2.79-3.95), immune and metabolic disorders (OR: 2.39, 95% CI: 2.00-2.85), respiratory diseases (OR: 2.02, 95% CI: 1.80-2.26), cerebrovascular diseases (OR: 4.12, 95% CI: 3.04-5.58), any types of cancers (OR: 2.22, 95% CI: 1.63-3.03), renal (OR: 3.02, 95% CI: 2.60-3.52), and liver diseases (OR: 1.44, 95% CI: 1.21-1.71).

**Conclusions:** This study provides evidence of a higher likelihood of deaths among COVID-19 patients against morbidity categories. These findings could potentially help healthcare providers to sort out the most endangered COVID-19 patients by comorbidities, take precautionary measures during hospitalization, assess susceptibility to death, and prioritize their treatment, which could potentially reduce the number of fatalities in COVID-19.

## Introduction

The coronavirus disease 2019 (COVID-19) caused by the SARS-CoV-2, a virus from the large coronavirus family, started in a seafood market in Wuhan, China, and is now a global pandemic. The virus is highly transmissible (reproductive number: 1.6-6.5, doubling time: 6.4 to 7.4 days) [1], which can mainly transmit through respiratory droplets (coughs or sneezes), close contact with the infected person [2, 3], and touching surfaces or objects that are touched by the infected person [2, 3]. As of May 08, 2020, 130 days since the virus was first detected on December 31, 2019, approximately 3.77 million people from 215 countries or territories have been infected with this virus [4]. Around 0.26 million of them have already died [4], and about 2% of currently infected people are now in critical conditions [5]. To date, there is no specific medicine or vaccine for COVID-19; therefore, the majority of the affected countries are taking non-pharmaceutical interventions such as restriction in inhabitants’ mobility, quarantine of suspected persons, isolation of infected persons, travel restrictions, and airport screening to reduce further infections [1, 6, 7].

The virus is equally transmissible in all ages; however, people who are now in critical conditions or who died were more likely to be in older age and found they had one or more morbidities [8-11]. Commonly reported morbidities among patients who died from COVID-19 were hypertension, diabetes, cardiovascular disease, and cerebrovascular disease [8, 12-14]. Notably, these comorbidities are independent causes of millions of annual deaths globally; 17.9 million deaths from cardiovascular system diseases, 9 million deaths from cancers, 3.9 million deaths from respiratory diseases, and 1.6 million deaths from diabetes, according to a report by World Health Organization (WHO) in 2018 [15]. People with one or more of these morbidities usually have poor immune systems, which increases their susceptibility to being infected, to reach in critical condition, and even died from a secondary disease like COVID-19 [1, 12, 16-19]. Precautionary measures following COVID-19 among patients with one or more morbidities could be potential ways to combat its adverse outcomes and severities. Thus, we need to identify possible morbidities that are potentially increasing the risks of mortality, which are still lacking. Studies conducted among COVID-19 patients are highly varied with reported morbidities and the likelihood of mortality [8, 10, 20, 21]. To address these gaps, this study was conducted with two primary aims: (i) to summarize pre-existing morbidities in patients with COVID-19, a secondary disease, and (ii) to estimate the likelihood of mortality from COVID-19 against each category of pre-existing morbidities. The study findings could help healthcare providers to take appropriate measures to control fatalities from this pandemic.

## Methods

This systematic review and meta-analysis was conducted by following the Preferred Reporting for Systematic Review and Meta-Analysis (PRISMA) consensus statement [22]. Studies relevant to the COVID-19 disease among people with pre-existing one or more morbidities were included.

### Search strategy

Four databases: Medline, Web of Science, Scopus, and CINAHL were searched, concluded on May 01, 2020, using pre-specified search strategies for each database. The search strategy consists of keywords on COVID-19 disease (COVID-19, 2019-nCoV, Coronavirus, SARS-CoV-2), pre-existing morbidity (comorbidity, morbidity), and patients’ survival status (mortality, death, died) combined using the Boolean operators (AND, OR). Details of the search strategies are presented in the supplementary tables (Table 1-4). Additional searches were conducted using the reference list of the selected studies, relevant journal websites, and renowned pre-print servers (medRxiv, bioRxiv, SSRN).

**Table 1.**
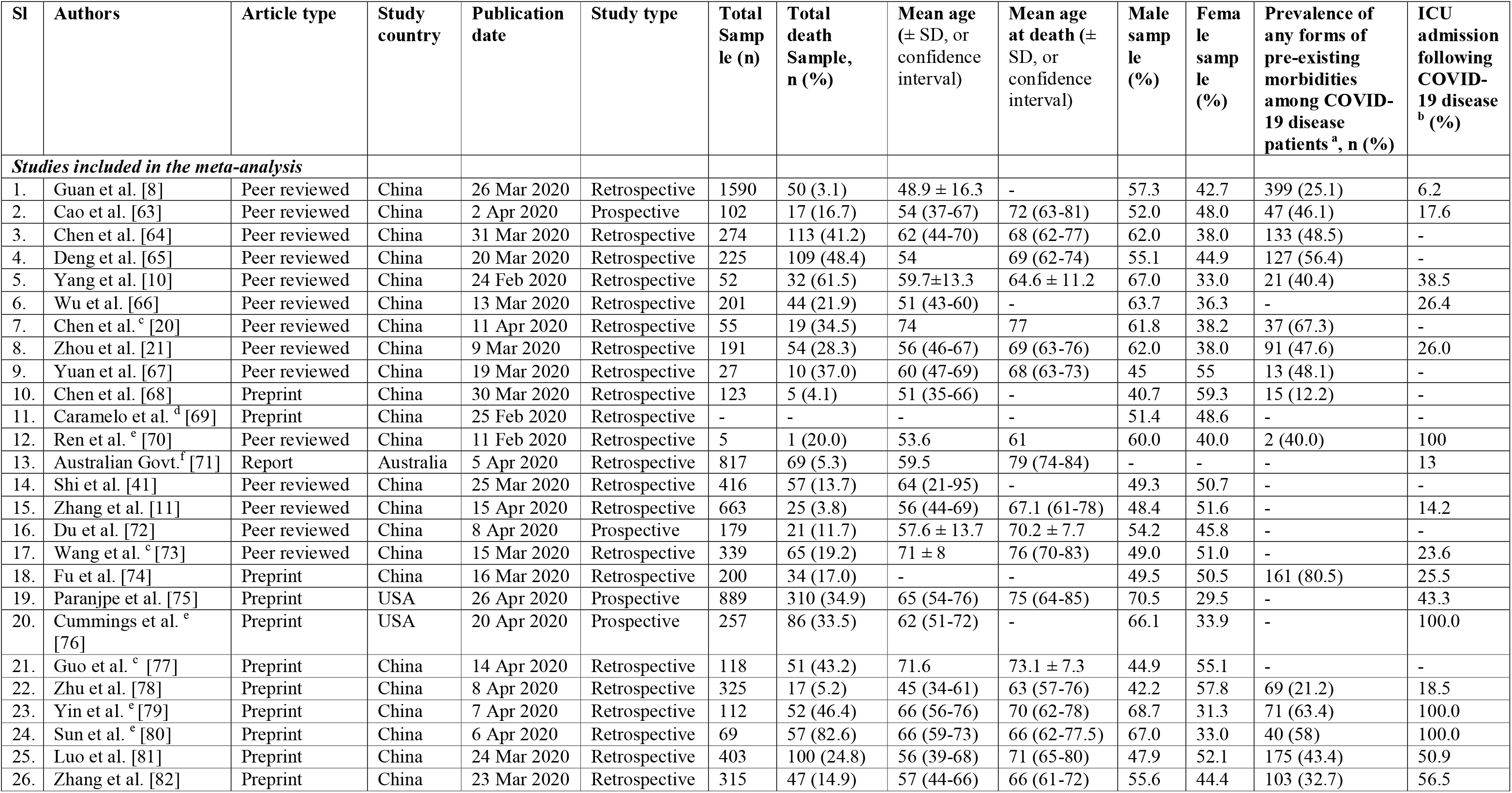
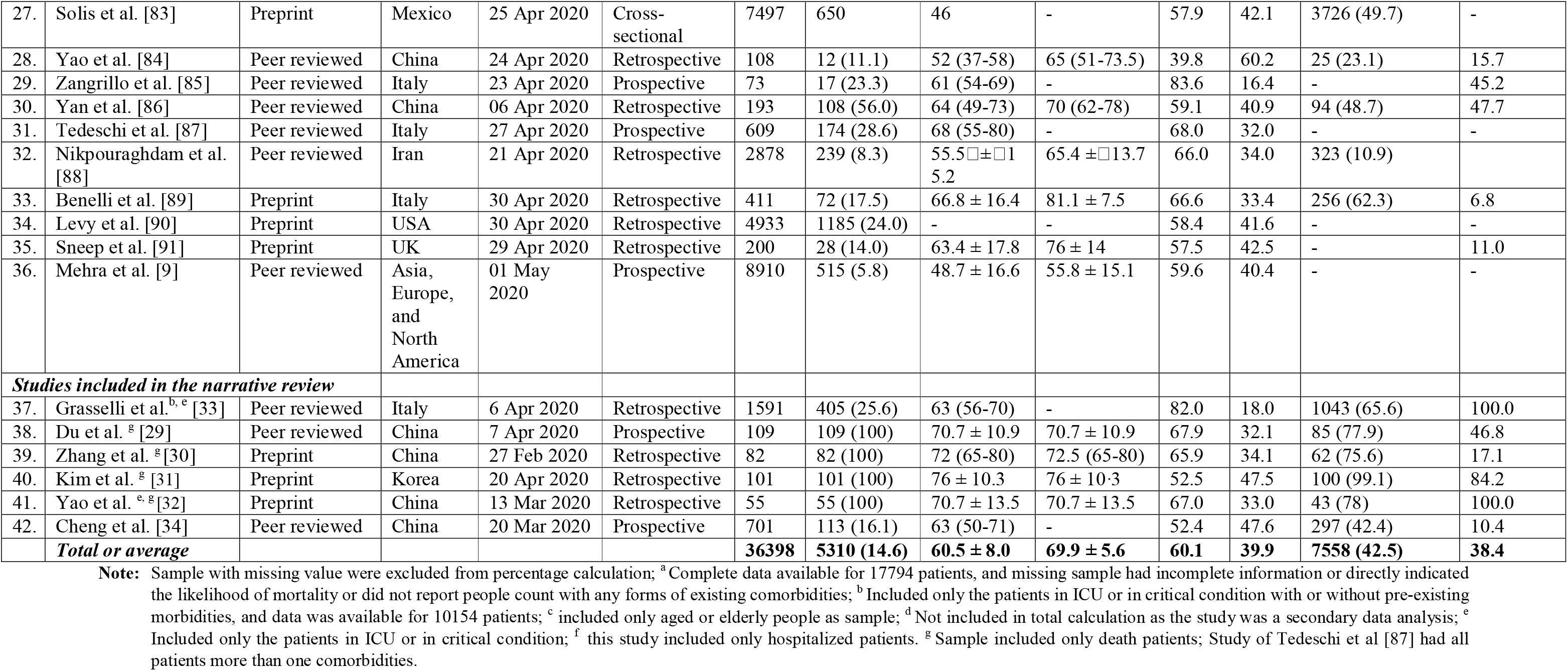
Background characteristics of the selected studies and hospitalized sample (n = 36,398).

**Table 2.** Percentage distribution of comorbidities among patients reported in admission COVID-19 infection.

| **Pre-existing morbidities** | **Distribution of comorbidities for total patients** | **(%)** |
| --- | --- | --- |
| ***Cardiovascular system disease*** | **12760** | **51.2** |
| Hypertension | 9085 | 36.5 |
| Cardiovascular Disease <sup>a</sup> | 2957 | 11.9 |
| Arrhythmia | 393 | 1.6 |
| Heart failure | 325 | 1.3 |
| ***Immune and metabolic disorders*** | **5966** | **23.9** |
| Diabetes | 5478 | 22.0 |
| Immunosuppression | 406 | 1.6 |
| Autoimmune disease | 8 | 0.03 |
| Immunodeficiency | 3 | 0.01 |
| Metabolic disorder | 4 | 0.02 |
| ***Respiratory system diseases*** | **1975** | **7.9** |
| Chronic lung disease | 1010 | 4.1 |
| Chronic obstructive pulmonary disease (COPD) | 577 | 2.3 |
| Asthma | 355 | 1.4 |
| Acute respiratory distress syndrome (ARDS) | 11 | 0.04 |
| Chronic bronchitis | 10 | 0.04 |
| Tuberculosis | 9 | 0.04 |
| Pulmonary emphysema | 3 | 0.01 |
| ***Any types of cancer*** | **224** | **0.9** |
| Malignancy | 33 | 0.1 |
| Cancer | 92 | 0.4 |
| Tumor | 16 | 0.1 |
| Carcinoma | 4 | 0.02 |
| ***Cerebrovascular system diseases*** | **198** | **0.8** |
| Cerebrovascular disease <sup>b</sup> | 189 | 0.8 |
| Peripheral vascular disease | 9 | 0.04 |
| ***Renal system diseases*** | **779** | **3.1** |
| Chronic kidney disease | 758 | 3.0 |
| Urinary disease | 21 | 0.1 |
| ***Liver system diseases*** | **2844** | **11.4** |
| Chronic liver disease | 45 | 0.2 |
| Cirrhosis | 25 | 0.1 |
| Fatty liver disease | 15 | 0.1 |
| Hepatitis B | 21 | 0.1 |
| Hyperlipidemia | 2732 | 11.0 |
| Inflammatory disease | 6 | 0.02 |
| ***Gastrointestinal system diseases*** | **61** | **0.2** |
| Chronic digestive disorder | 21 | 0.1 |
| Gastrointestinal disease | 40 | 0.2 |
| **Others <sup>c</sup>** | **109** | **0.4** |
| **Grand total** | **24916** | **100.0** |

**Note:** Patients with more than one comorbidity were missing. Calculated in column percentage format; One study (Caramelo et al.) was excluded from prevalence calculation as the study had not reported frequency of comorbidities; Malnutrition and dementia was skipped from the analysis as we found only one patient (Yang et al.); <sup>a</sup> Included all types of cardiovascular diseases like coronary heart diseases or artery disease; <sup>b</sup> Included Cerebral infarction; <sup>c</sup> anemia, bowel disease, tissue disease, etc.

**Table 3.** Summary effects of type of morbidity categories on death among patients infected with COVID-19, publication bias, and Trim and Fill

| Characteristics | Number of studies | Number of times morbidity reported | Summary estimates |  | Egger bias test p-value | Trim and Fill estimates <sup>a</sup> |  |
| --- | --- | --- | --- | --- | --- | --- | --- |
| | | | OR (95% CI) | Heterogeneity Index ($I^2$) | | Missing studies no. | OR (95% CI) |
| Cardiovascular system diseases | 33 | 64 | 3.32 (2.79-3.95) <sup>b</sup> | 83.8% | 0.001 | 13 | 2.82 (2.38-3.34) |
| Immune and metabolic disorders | 31 | 38 | 2.39 (2.00-2.85) <sup>b</sup> | 64.5% | 0.019 | 6 | 2.14(1.78-2.57) |
| Respiratory system diseases | 28 | 33 | 2.02 (1.80-2.26) <sup>c</sup> | 71.2% | 0.031 | 3 | 2.36 (1.79-3.11) |
| Any types of cancer | 20 | 20 | 2.22 (1.63-3.03) <sup>c</sup> | 67.7% | 0.891 | 0 | 2.22 (1.63-3.03) |
| Cerebrovascular system diseases | 15 | 16 | 4.12 (3.04-5.58) <sup>c</sup> | 25.7% | 0.048 | 2 | 3.94 (2.92-5.31) |
| Renal system diseases | 21 | 21 | 3.02 (2.60-3.51) <sup>c</sup> | 56.0% | 0.024 | 4 | 2.86 (2.47-3.32) |
| Liver system diseases | 14 | 17 | 1.44 (1.21-1.71) <sup>c</sup> | 0.0% | 0.001 | 6 | 1.38 (1.16-1.63) |
| Gastrointestinal system diseases | 5 | 5 | 1.33 (0.56-3.19) <sup>c</sup> | 0.0% | 0.170 | 0 | 1.33 (0.56-3.19) |

estimates
**Note:** Person survived from COVID-19 infection is considered as reference category; CI, confidence interval; OR, odds ratio.
<sup>a</sup> The trim-and-fill method simulates studies that are likely to be missing from the literature due to publication or other forms of bias. The trim-and-fill OR estimates what the pooled OR would be if these missing studies were included in the analysis; <sup>b</sup> Summary estimates were based on fixed-effects methods; <sup>c</sup> Summary estimates were based on random-effects methods.

**Table 4.** Stratified analysis of the likelihood of death among patients with cardiovascular system diseases infected with COVID-19

| Characteristics | Pooled OR (95% CI) | P |  |
| --- | --- | --- | --- |
|  |  | Heterogeneity | Meta-regression |
| **Type of diseases** |  |  |  |
| Cardiovascular diseases | 3.35 (2.53-4.42) | <0.01 | 0.95 |
| Hypertension | 3.28 (2.53-4.24) | <0.01 |  |
| Heart failure | 3.98 (2.96-5.34) | 0.348 |  |
| Arrhythmia | 3.00 (1.96-4.57) | 0.192 |  |
| **Study country** |  |  |  |
| Australia | 3.68 (2.06-6.57)<sup>++</sup> | NA | <0.01 |
| Italy | 3.95 (2.82-5.54) | **0.239** |  |
| China | 4.36 (3.40-5.58) | <0.01 |  |
| United State of America | 2.74 (2.00-3.75) | <0.01 |  |
| Asia, Europe, and North America | 1.98 (1.16-3.38) | <0.01 |  |
| Iran | 1.75 (0.95-3.22) | 0.685 |  |
| UK | 2.73 (1.17-6.35)<sup>++</sup> | NA |  |
| Mexico | 1.12 (0.71-1.75) | <0.05 |  |
| **Study design** |  |  |  |
| Cross-sectional | 3.79 (2.86-5.03) | <0.01 | <0.05 |
| Prospective cohort | 3.15 (2.25-4.41) | <0.01 |  |
| Retrospective cohort | 1.82 (0.71-1.75) | <0.05 |  |
| **Adjustment factor** |  |  |  |
| Unadjusted | 3.94 (3.24-4.80) | <0.01 | <0.05 |
| Adjusted | 1.81 (1.30-2.53) | <0.01 |  |
| **Sample size** |  |  |  |
| >1134 | 2.19 (1.59-3.02) | <0.01 | <0.01 |
| ≤1134 | 3.78 (3.14-4.56) | <0.01 |  |
| **Mean age of total sample** |  |  |  |
| ≤60 | 3.70 (2.70-5.07) | <0.01 | 0.593 |
| >60 | 3.14 (2.60-3.80) | <0.01 |  |
| **Mean age of death sample** |  |  |  |
| ≤69 | 4.39 (3.05-6.32) | <0.01 | 0.100 |
| >69 | 3.00 (2.46-3.66) | <0.01 |  |

### Study selection criteria

All peer-reviewed and pre-print (not-peer-reviewed) studies met the pre-specified inclusion, and exclusion criteria were included in this study.

### Inclusion criteria

Studies met the following inclusion criteria were included if: (i) conducted for the patients infected with COVID-19 with or without pre-existing morbidities, (ii) presented survivor and non-survivor counts following COVID-19 among patients with or without preexisting morbidity or presented hazard/risk/odds ratio of deaths or survival following COVID-19 against the types of morbidities, and (iii) published in the English language. Studies without complete information but met our inclusion criteria were included in the narrative review.

### Exclusion criteria

Studies excluded if COVID-19 was reported among pregnant women or children (aged <18 years) and written in languages other than English. We also excluded review papers, correspondence, viewpoints, editorials, commentaries, and studies where no information related to the previous morbidity was reported.

### Data extraction and quality assessment

A data extraction form was designed, trialed, and modified to extract information from the selected studies. Two authors (MMAK and MGM) used the pre-designed form to extract information independently. The following information was extracted: study location, design, sample size, study population characteristics (e.g., age, gender), and survivor vs. non-survivor counts among COVID-19 patients with or without specific morbidity. If available, the odds/risk/hazard ratio of deaths among COVID-19 patients with comorbidities were extracted against the types of morbidity. Disagreements reported in data extraction were reviewed and solved by the corresponding and senior authors (MNK and MIK). The modified Newcastle-Ottawa scale, as part of the data extraction strategy, was used to assess the quality of selected studies.

### Statistical analysis

Pre-existing one or more morbidities among COVID-19 patients reported in the selected studies were grouped into eight broad categories based on the type of morbidities. These were cardiovascular system diseases (hypertension, cardiovascular disease, arrhythmia, heart failure), immune and metabolic disorders (diabetes, immunosuppression, autoimmune disease, immunodeficiency, metabolic disorder), respiratory system diseases (chronic lung diseases, Chronic Obstructive Pulmonary Disease (COPD), acute respiratory distress syndrome, tuberculosis, etc.), cancer (malignancy, cancer, and tumor), cerebrovascular diseases (cerebrovascular disease, peripheral vascular disease), renal system diseases (chronic kidney disease, urinary disease), liver system diseases (chronic liver disease, cirrhosis, hyperlipidemia, Hepatitis B, etc.), and gastrointestinal system diseases (chronic digestive disorder, gastrointestinal disease). The odds ratios (ORs) of deaths with 95% confidence interval (95% CI) for the people exposed to a particular category of morbidity as compared to people unexposed to any specific morbidity was estimated from the extracted raw data or reported ORs. We first used the Haldane correction (add constant 0.5 to each cell) for the studies in which the sample included in the exposed or unexposed group was zero (such as all exposed patients died or vice versa) [23-25]. We then used either a fixed effect or random effect model to estimate ORs, selected based on heterogeneity assessment. When the test of heterogeneity *(I^2^* statistics) was moderate (50-74%) or high (≥75%), the pooled estimates of ORs were computed using the random-effects model [26]. Subgroup and meta-regression analyses were conducted for the groups where moderate or higher heterogeneity was reported. For this, pre-specified subgroups (types of morbidities, study country, study design, mean age of the total sample, mean age of death sample) were used. Publication bias was assessed by visual inspection of the funnel plot asymmetry and Egger’s regression test [27]. When evidence of publication bias was found, the Trim and Fill method was used to estimate and adjust potentially missing studies, and the effect size was recalculated accordingly [28]. Stata software version 15.1 (StataCorp. LP, College Station, TX, USA) was used for all analyses.

## Results

A total of 247 articles were identified from the databases searched, and the additional 15 articles were identified by checking the reference list of the selected articles and the selected journal’s websites (Figure 1). Around 1273 articles were also initially identified from the aforementioned pre-prints servers. Of the selected articles, 1341 articles were excluded after screening titles and abstracts, leaving 114 articles for full-text review for possible inclusion in this study. Of these, 55 articles were excluded based on the inclusion and exclusion criteria for the study sample (e.g., excluded pregnant or children), and 11 articles were excluded for study types (e.g., review papers, correspondence, viewpoints, editorials, commentaries), and six articles were excluded for entirely incomplete data. A total of 42 articles were finally selected for this study; 36 articles were included in the meta-analysis, and the remaining six articles were synthesized narratively.

**Figure 1.**
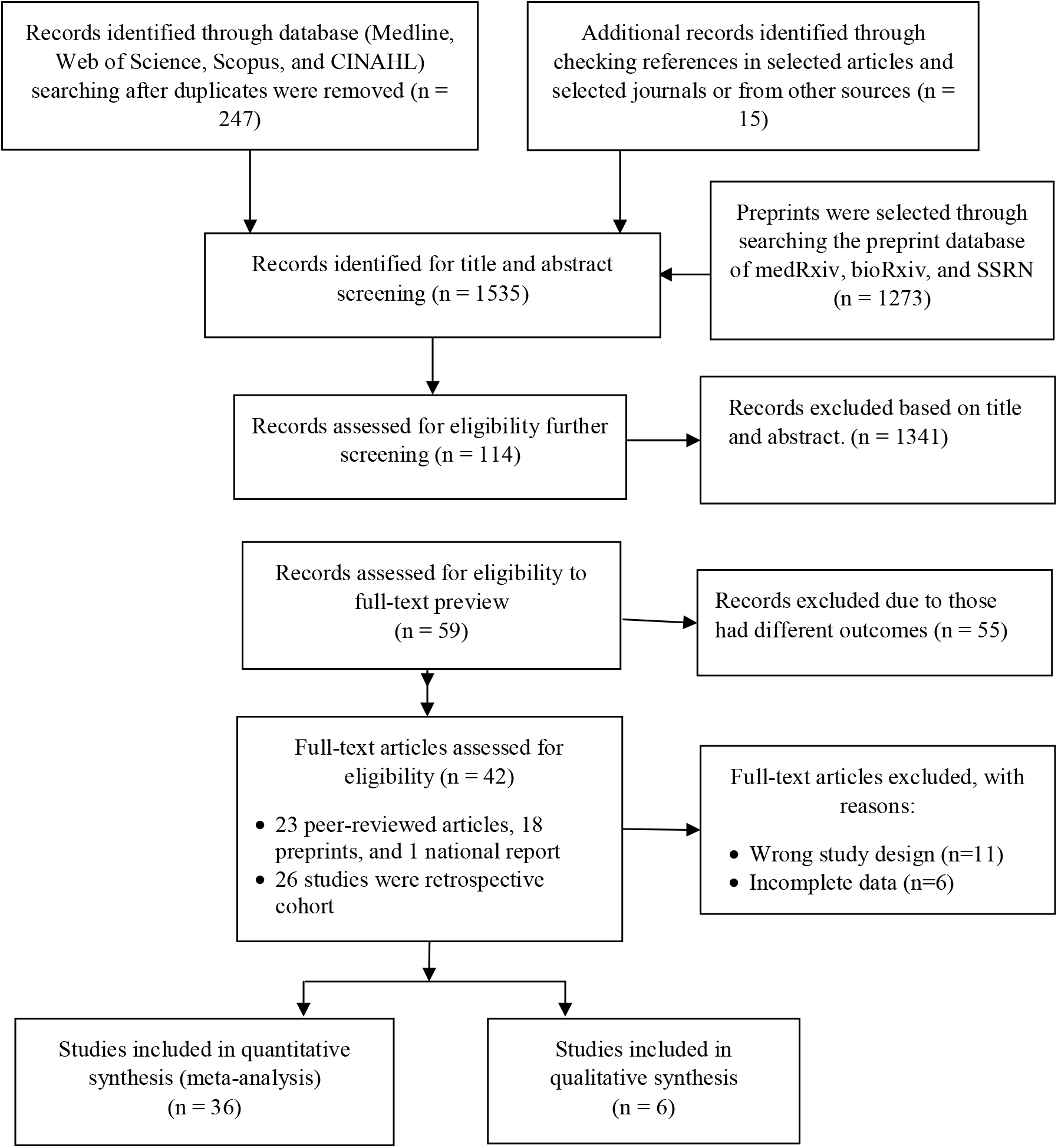
Schematic representation of studies included in the systematic review and meta-analysis using PRISMA checklist and flow diagram

### Study characteristics

A summary of the 42 selected articles is represented in Table 1. A total of 23 of the selected 42 articles were published in peer-reviewed journals, and 18 articles were published in pre-print servers. One of the selected studies was a national report for Australia. The majority of these studies were retrospective in nature (26), along with seven prospective studies. The sleeted studies comprised 36,398 COVID-19 patients, 7,558 (42.5%) of them had preexisting one or more morbidities, 38.4% of patients had undergone critical care, and 5,310 (14.6%) of them died. Their average age was 60.5 ± 8.0years, and 60.1% of them were male. The mean age at death was 69.9 ± 5.6 for the patients who died in COVID-19. A total of 36 selected studies presented death counts following COVID-19 among patients with or without specific one or more morbidities. Four included studies (Du et al. [29], Zhang et al. [30], Kim et al. [31], and Yao et al. [32]) were conducted only for dead COVID-19 patients and reported the status of pre-existing morbidities before their deaths. All studies were of moderate to high quality (Supplementary Table 6-7).

### Prevalence of pre-existing morbidity among COVID-19 patients

Distribution of the type of morbidity presented in Table 2. Approximately 36.5% of the total COVID-19 patients reported that they had hypertension, 22.0% had diabetes, 11.9% had cardiovascular disease, 4.1% had chronic lung disease, 2.3% had COPD, 11.0 % had hyperlipidemia, and 3.0% had chronic kidney disease.

### Effects of preexisting morbidity on deaths in COVID-19 patients

The pooled ORs of deaths for each category of pre-existing morbidities among COVID-19 patients, publication bias, and Trim and Fill estimates are presented in Table 3. COVID-19 patients with preexisting cardiovascular system disease were 3.32 times more likely to die (OR: 3.32, 95% CI: 2.79-3.95; *I^2^*= 83.8%) than the patients who had no cardiovascular system diseases. The odds of death among COVID-19 patients with immune and metabolic disorders were also found to be 239% higher (OR: 2.39, 95% CI: 2.00-2.85; *I^2^*= 64.5%) than among COVID-19 patients without such disorders. The incidence of COVID-19 among people with respiratory system disease increases mortality risk around two times (OR: 2.02, 95% CI: 1.80-2.26; *I^2^*= 71.2%) than COVID-19 patients without respiratory system diseases. Similarly, we found higher mortality risk among COVID-19 patients who had pre-existing any types of cancers (OR: 2.22, 95% CI: 1.63-3.03, *I^2^=* 67.7%) and cerebrovascular system diseases (OR: 4.12, 95% CI: 3.04-5.58) than their counterparts. Moreover, the incidence of COVID-19 among patients with pre-existing renal system disease and chronic liver disease increased mortality risk by about three times (OR: 3.02, 95% CI: 2.60-3.51) and one and half times (OR: 1.44, 95% CI: 1.21-1.71), respectively compared to the COVID-19 patients who did not mention such comorbidities.

We found evidence of publication bias for the three categories of pre-existing morbidities: any type of cancer, cerebrovascular diseases, and liver system diseases (Figure 1a to 8a). We then used Trim and Fill methods to impute the number of missing studies, which hypothetically imputed two studies for cardiovascular system diseases, three studies for renal system diseases, and three studies for liver system diseases. The pooled analysis, including these missing studies, showed almost similar results to the summary estimates presented earlier without these missing studies.

Evidence of higher deaths among COVID-19 patients with preexisting one or more morbidities were also demonstrated in the narrative review (Supplementary Table 5). In two of the three articles reviewed in this study, researchers reported that each of the patients who died following COVID-19 had pre-existing morbidities, mostly had any types of cardiovascular diseases and immune and metabolic disorders [29, 30]. Researchers in one study found around 38% of the COVID-19 patients with hypertension died [33], and one study reported higher odds for deaths in kidney injury [34].

### Stratified analysis

We found evidence of high heterogeneity (I^2^ > 75%) for cardiovascular system diseases (83.8%). To examine the sources of heterogeneity, we conducted stratified analysis across types of comorbidities, study design (cross-sectional vs. retrospective cohort vs. prospective cohort), sample size (divided based on mean sample size of the included studies and classified as ≤ 1134, > 1134), age of the total sample (divided based on mean age and classified as ≤60 years and >60 years), and age at death (divided based on mean age and classified as ≤69 vs.>69) (Table 4). We found odds of death varied across specific types of pre-existing morbidities included to generate the cardiovascular system diseases category. For instance, in the cardiovascular system diseases category, the odds of mortality were found higher for COVID-19 patients with pre-existing heart failure (OR: 3.98, 95% CI: 2.96-5.35), cardiovascular disease (OR: 3.35, 95% CI: 2.53-4.42), hypertension (OR: 3.28, 95% CI: 2.53-4.24), than for COVID-19 patients with pre-existing arrhythmia (OR: 3.00, 95% CI: 1.96-4.57.

## Discussion

This study aimed to summarize pre-existing morbidities among COVID-19 patients, which increases their incidence of deaths and their corresponding likelihoods. A total of 42 studies were included that comprised 36,398 samples, and 7,558 (42.5%) of them had pre-existing morbidities. The most frequently reported morbidities were hypertension (36.5%), diabetes (22.0%), and cardiovascular disease (11.9%). The likelihood of death was higher among COVID-19 patients who had comorbidities like cardiovascular and cerebrovascular diseases, respiratory diseases, renal diseases, immune and metabolic disorders, hepatic diseases, and cancer. This evidence will guide physicians to take precautionary measures, which could reduce the number of fatalities following secondary infection with COVID-19.

Among the total positive COVID-19 cases included in this systematic review, around 43% had pre-existing one or more morbidities, mostly cardiovascular diseases and immune and metabolic disorders. Importantly, patients with these diseases are more likely to have a higher neutrophil-lymphocyte ratio [35, 36], higher D-dimer level [37], and higher C-reactive protein [38]. These increased parameters lead to multiple organ failure [39, 40], severe pneumonia, hypoxia, respiratory failure, myocardial damage, and circulatory failure [18]. These non-communicable diseases independently elevate the risk of death and increase further if patients are infected with COVID-19 [21, 40-42]. COVID-19 also damages patients’ myocardial cells by destabilizing coronary plaque in pre-existing cardiovascular conditions and previous history of myocardial infarct [43, 44]. Similar higher risks of mortality were reported among SARS-CoV and MERS-CoV patients with cardiovascular diseases [41, 43, 45, 46]. These two diseases are considered as ancestors of current COVID-19, which were reported in 2003 and 2012, respectively [41, 43, 45, 46]. Evidence also validates that the occurrence of influenza, along with cardiovascular diseases and diabetes, could increase the risk of death [47, 48].

Pooled likelihoods in this study provide evidence of higher deaths among COVID-19 patients who had pre-existing chronic respiratory diseases or any type of cancers. Chronic respiratory diseases like COPD and asthma are well-established risk factors for pneumonia [49], which also increase the susceptibility to COVID-19 infection [50]. Once patients are infected with COVID-19, these further affect the patient’s respiratory system and progress to severe hypoxemia [51]; therefore, the cumulative effects lead to events of death [18, 50]. Cancer patients are more likely to report a systemic immunosuppressive state and progress to severe clinical events, such as require intensive care (ICU) or death [52, 53]. Secondary infection of COVID-19has its own adverse consequences on the human body, which could,therefore, increase serve clinical events as well as deaths among patients with these pre-existing morbidities.

This study also suggests that patients with cerebrovascular, liver, and renal diseases are more vulnerable to mortality following the second incidence of COVID-19 than the patient does not have such diseases. The results are comparable to deaths among previously reported SARS patients [54]. Comorbidities such as, cardio-cerebrovascular diseases, liver damage, or renal diseases accelerate an abrupt loss of kidney function [55, 56], tissue damage that causes hypoxia, shock, and rhabdomyolysis [34, 57], and increased occurrence of thrombocytopenia (reduced platelet counts) [21, 58]. These could independently elevate the risk of death and add to the adverse effects on the human body being infected with COVID-19. Together, these increase occurrences of deaths. Moreover, elevated alanine aminotransferase (ALT) levels and reduced albumin levels are found to be associated with higher mortality in COVID-19 [21, 58], which can be caused by chronic liver and kidney diseases [58-62]. Thus, it indicates an urgency of early precautions to reduce and prevent deaths among COVID-19 patients with pre-existing morbidities.

### Strengths and limitations

This study has several strengths and limitations that should be reported. To our knowledge, this is the first of its kind that summarizes all morbidities among COVID-19 patients that lead to death. Moreover, morbidities reported among COVID-19 patients were classified into board groups based on their characteristics, and the likelihood of death was estimated separately for each group. This evidence informs healthcare providers about the risk of death among COVID-19 patients with different groups of pre-existing morbidities. Thus, they will be able to take precautionary measures early targeting to prevent deaths. However, this study reported the odds of death for COVID-19 patients with one pre-existing morbidity only. Many COVID-19 patients may have multi-morbidities (COVID-19 with pre-existing two or more morbidities) and a higher risk of death. However, the studies included in this review considered each morbidity separately; for instance, if COVID-19 patients had both hypertension and diabetes, they were included in both groups. None of the included studies considered COVID-19 with two or more morbidities together; therefore, we failed to provide the likelihood of deaths for COVID-19 patients with two or more pre-existing morbidities.

Moreover, the likelihoods presented in this study were mostly unadjusted (31 of the 36 articles included) calculated from the extracted raw data. This may overestimate or underestimate the actual likelihood of deaths in COVID-19 patients because age and other socio-demographic characteristics are potential confounders of their deaths, which should be adjusted for getting unbiased estimates. Despite these limitations, this study is unique and beneficial for healthcare providers to handle COVID-19 patients with pre-existing morbidities.

## Conclusion

About 46% of the sample included in this systematic review had one or more pre-existing morbidities and got COVID-19 as a secondary infection. The most common pre-existing morbidities were hypertension, diabetes, and cardiovascular disease. The likelihood of death was higher among COVID-19 patients who had pre-existing cardiovascular and cerebrovascular diseases, respiratory diseases, renal diseases, immune and metabolic disorders, liver diseases, and any types of cancer. These findings will help healthcare providers to sort COVID-19 patients by comorbidities, take precautionary measures during hospitalization, assess susceptibility to death, and prioritize their treatment. These could potentially reduce the number of fatalities from secondary infection with COVID-19 disease.

## Data Availability

Data related to this study are available upon request to the corresponding author.

---

**Supplementary Figure 1:**
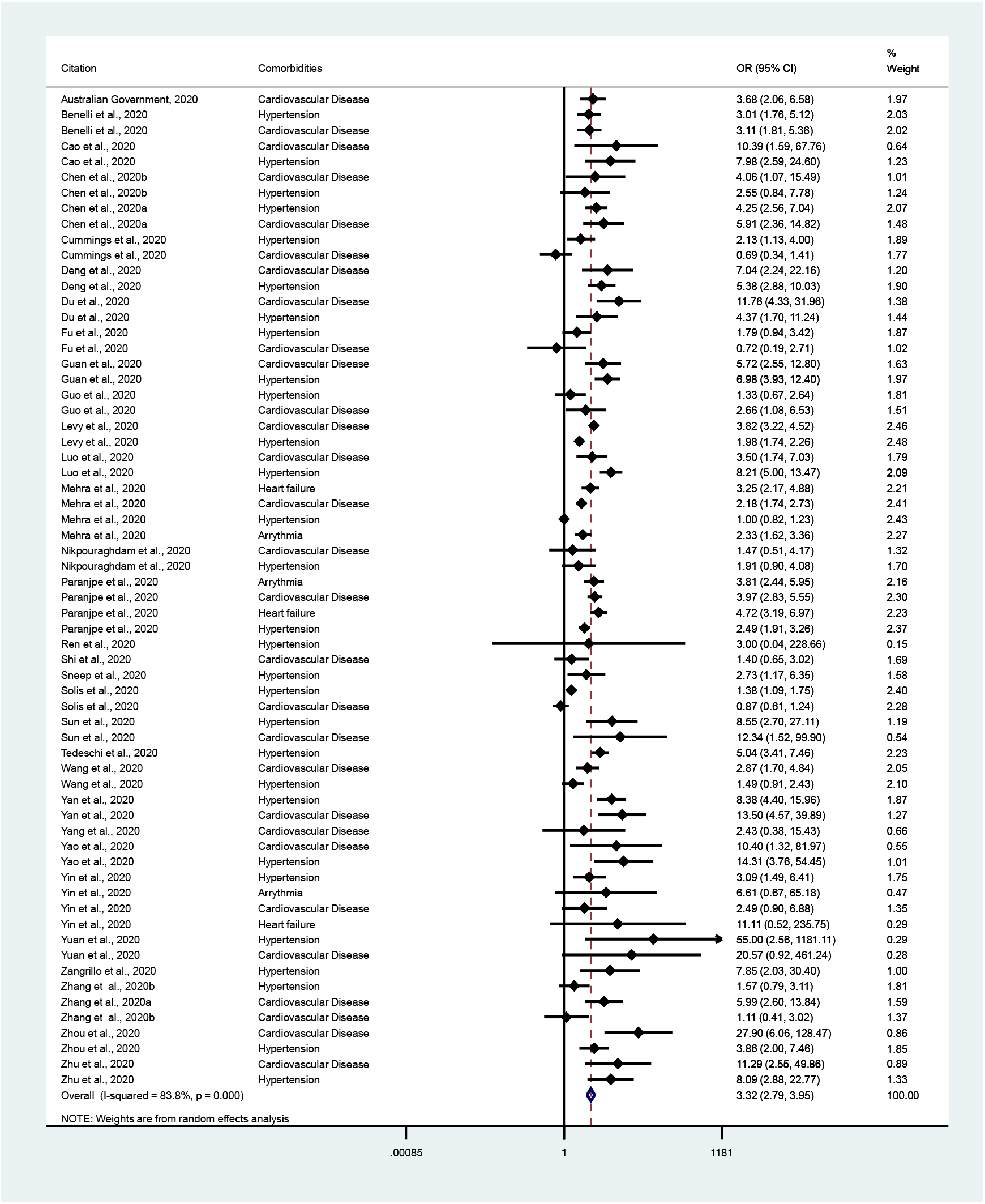
Likelihoods of death among patients with cardiovascular system diseases infected further with COVID-19 disease

**Supplementary Figure 1a:**
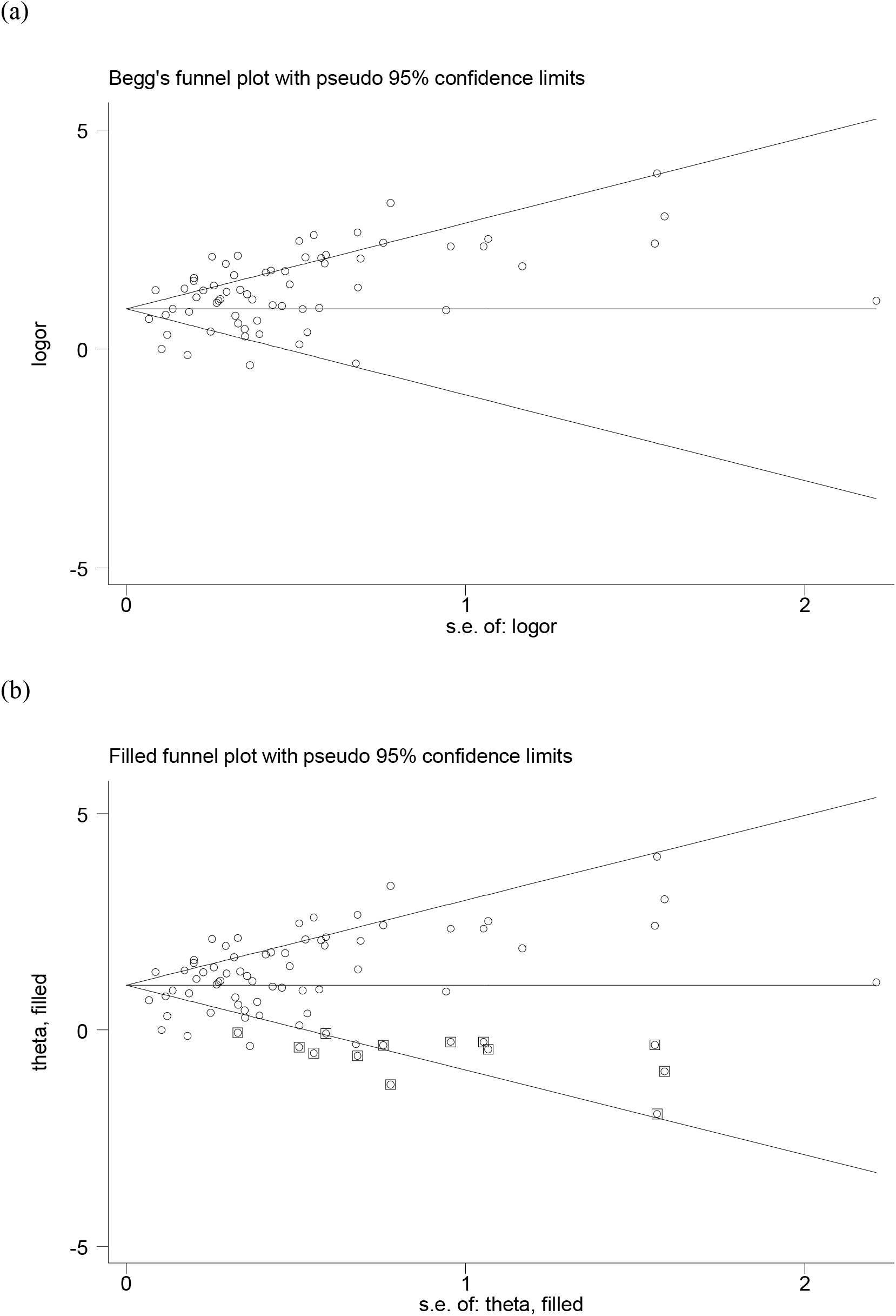
Funnel plot without (a) and with trim and fill (b) estimate for cardiovascular systems diseases patients infected further with COVID-19 disease

**Supplementary Figure 2:**
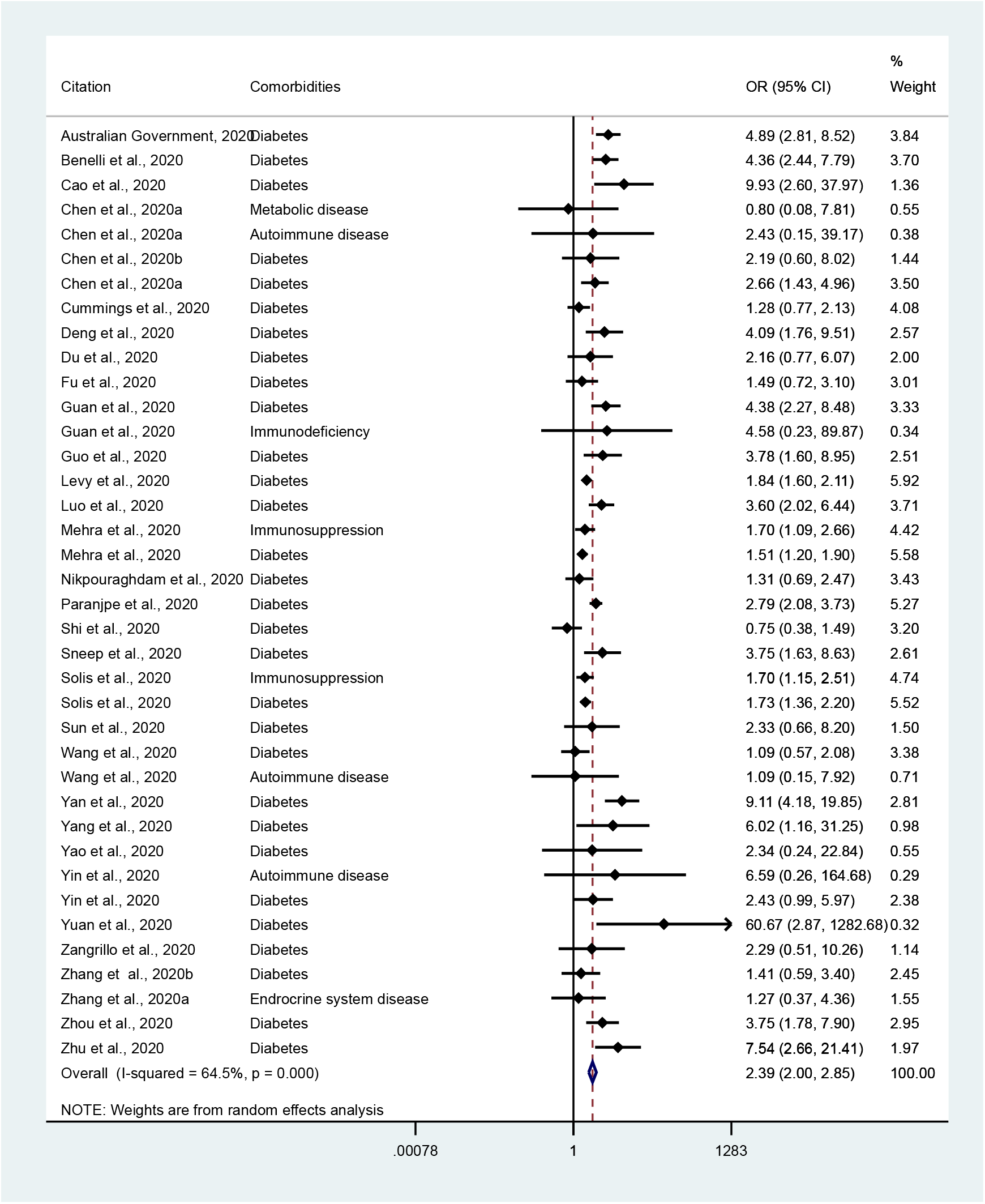
Likelihoods of death among patients with Immune and metabolic disorders patients infected further with COVID-19 disease

**Supplementary Figure 2a:**
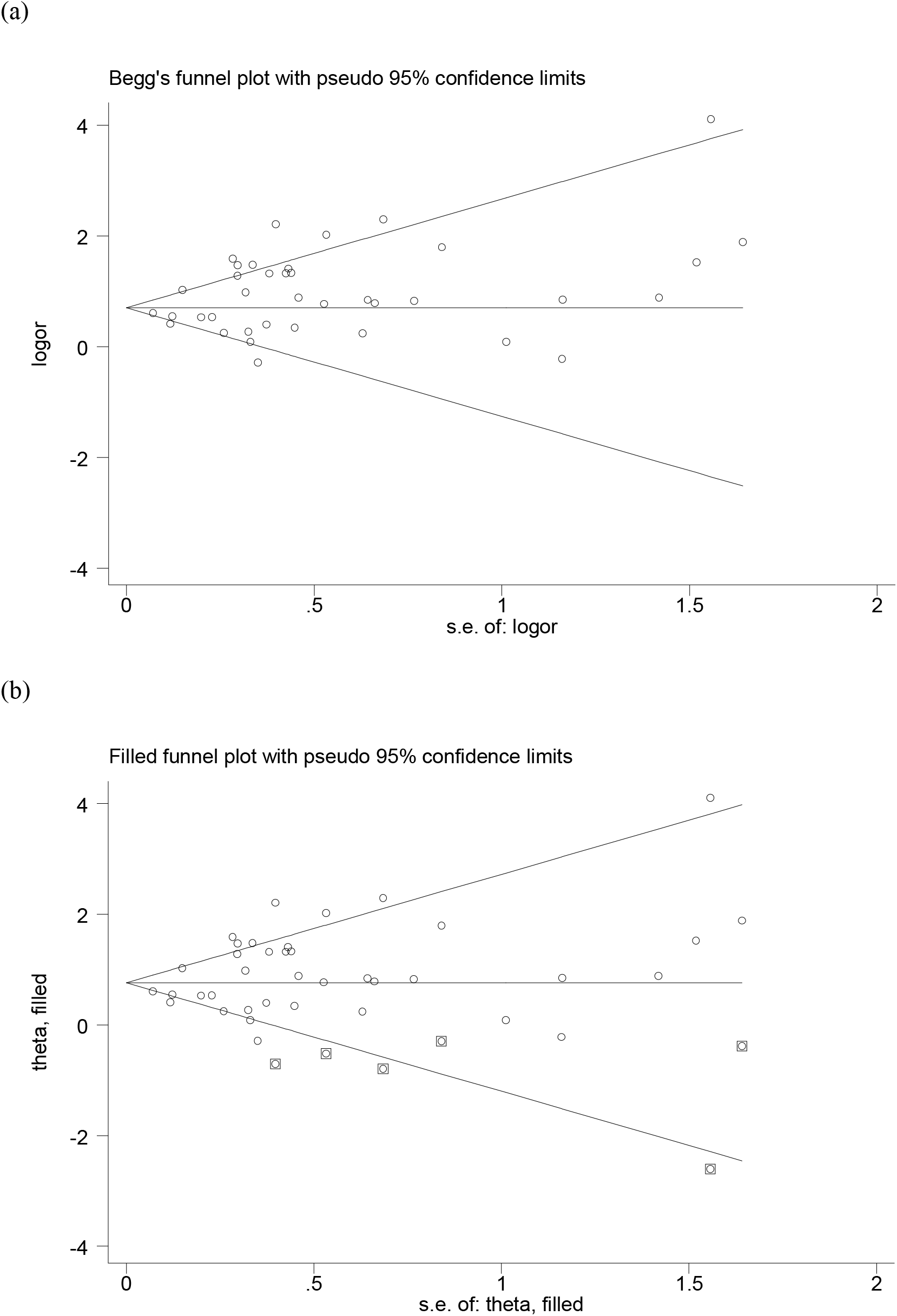
Funnel plot without (a) and with trim and fill (b) estimate for Immune and metabolic disorders patients infected further with COVID-19 disease

**Supplementary Figure 3:**
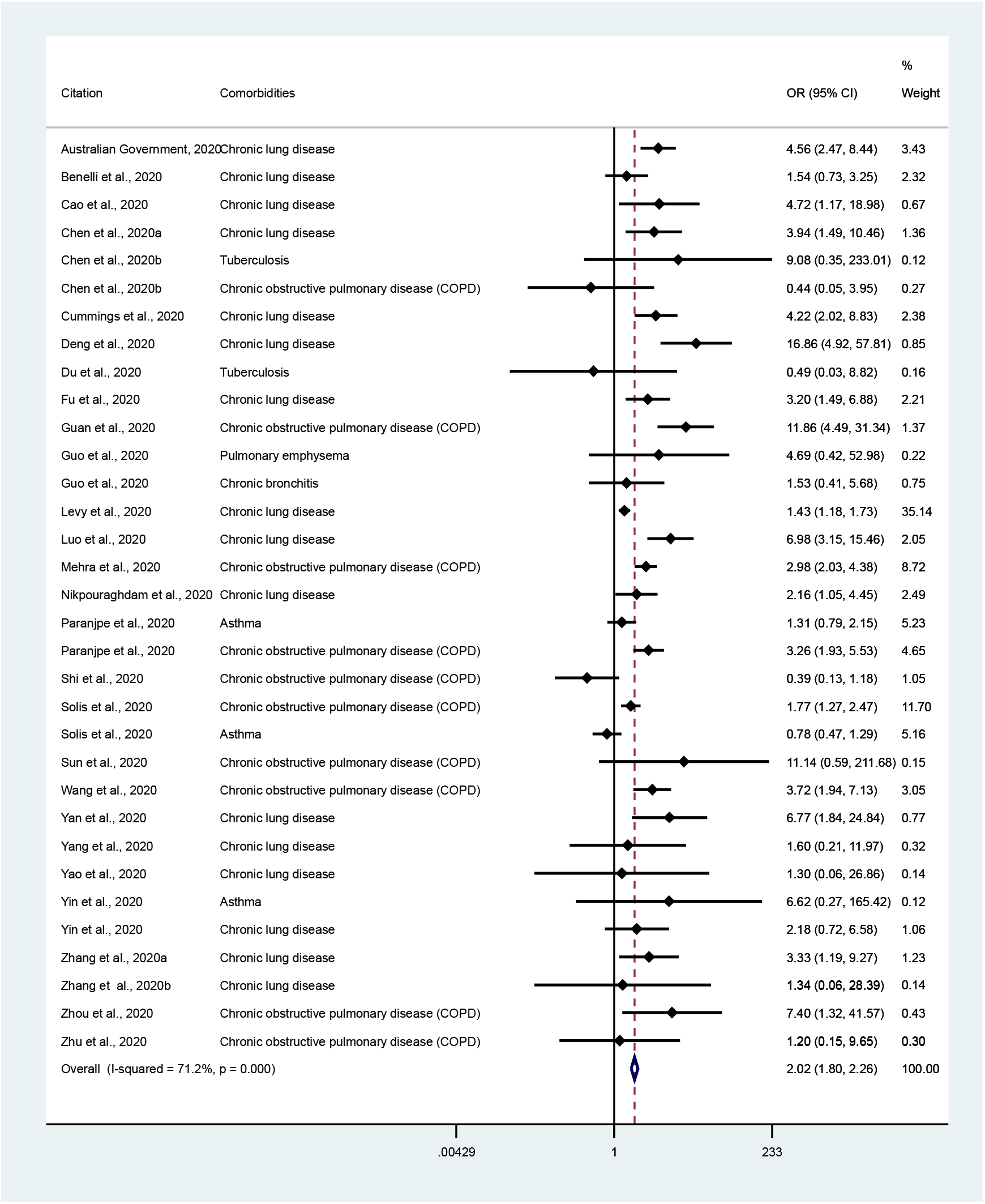
Likelihoods of death among patients with respiratory system diseases patients infected further with COVID-19 disease

**Supplementary Figure 3a:**
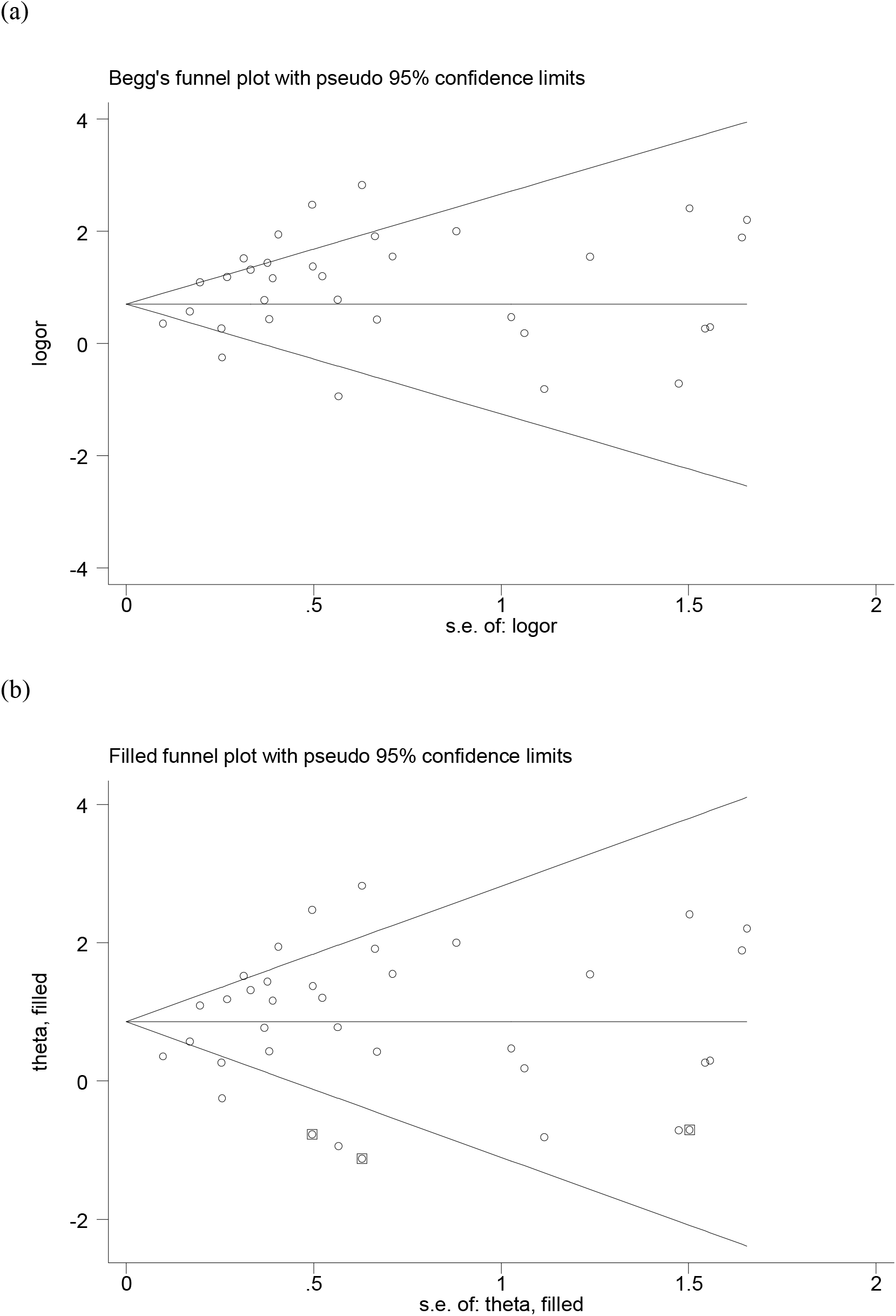
Funnel plot without (a) and with trim and fill (b) estimate for Respiratory system diseases patients infected further with COVID-19 disease

**Supplementary Figure 4:**
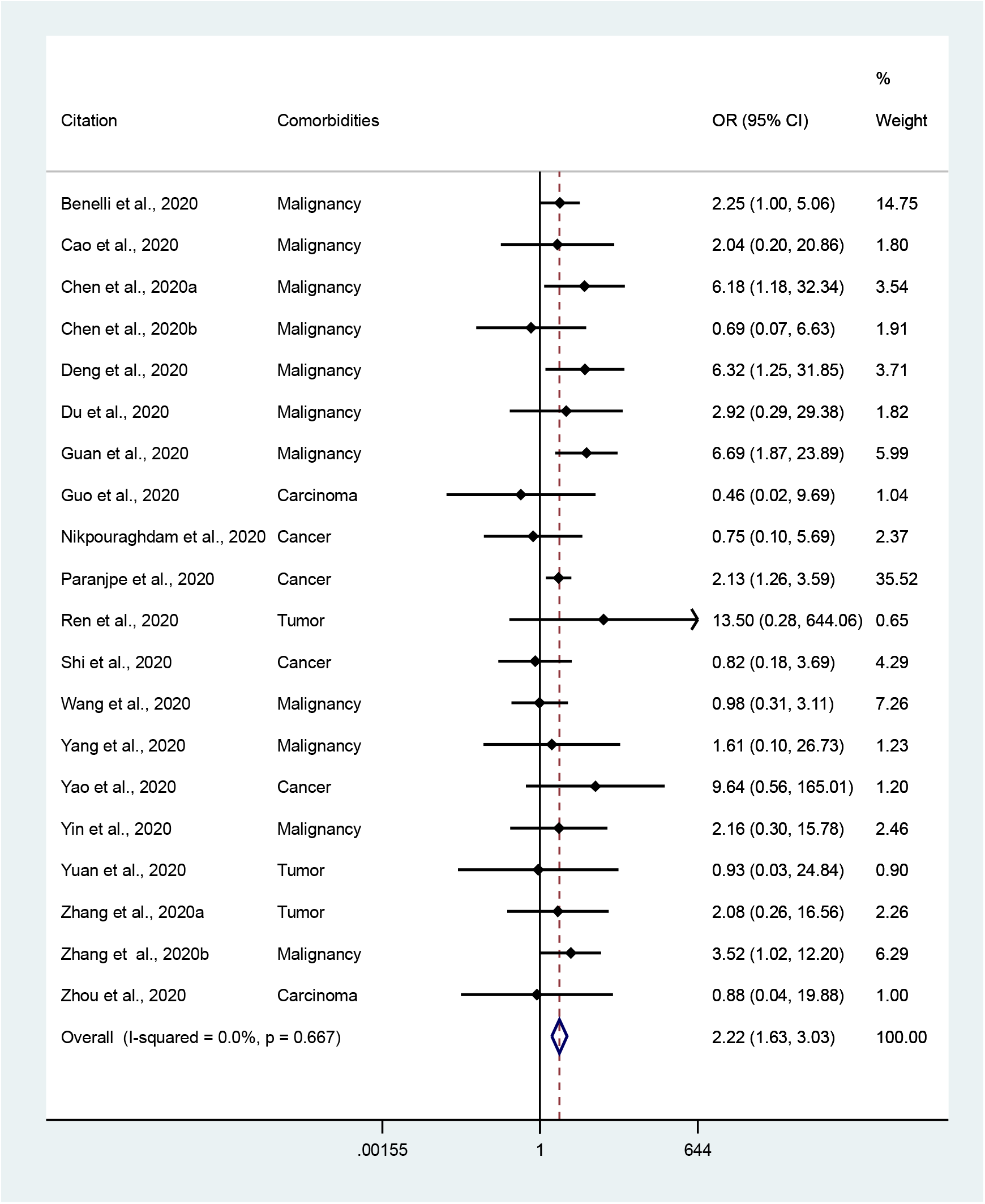
Likelihoods of death among patients with any type of cancers infected further with COVID-19 disease

**Supplementary Figure 4A:**
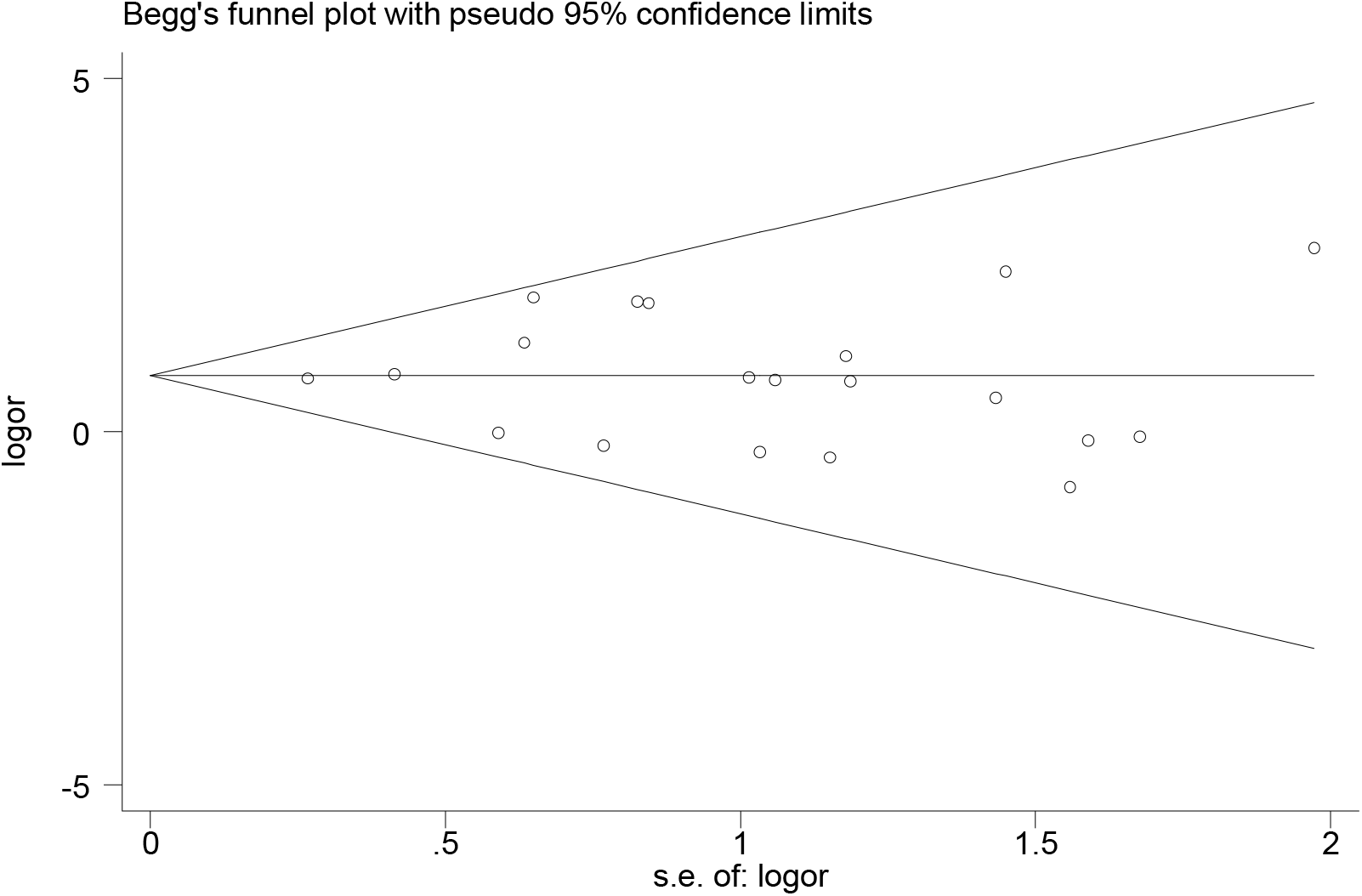
Funnel plot for patients with any type of cancers infected further with COVID-19 disease

**Supplementary Figure 5:**
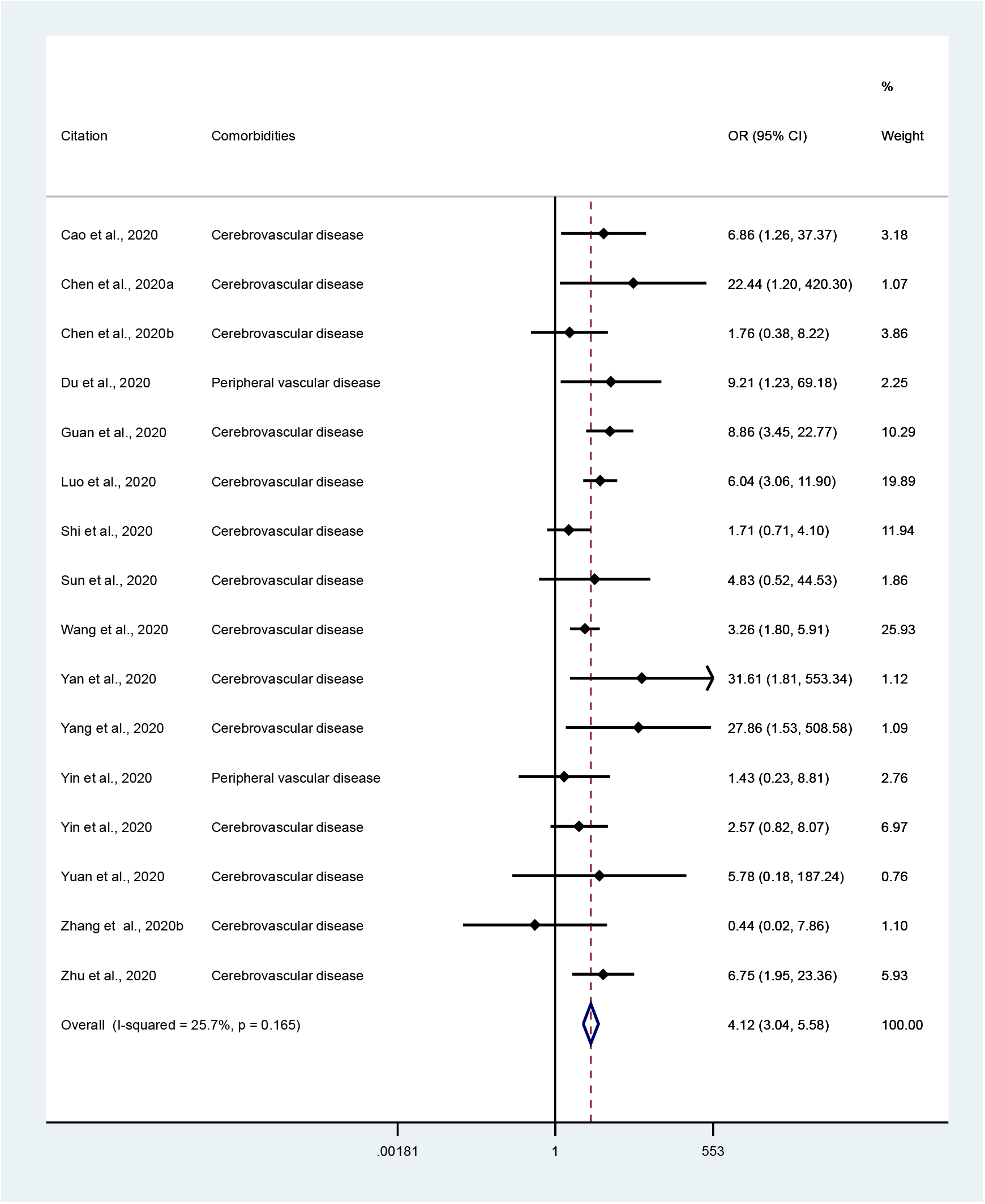
Likelihoods of death among patients with cerebrovascular system diseases patients infected further with COVID-19 disease

**Supplementary Figure 5A:**
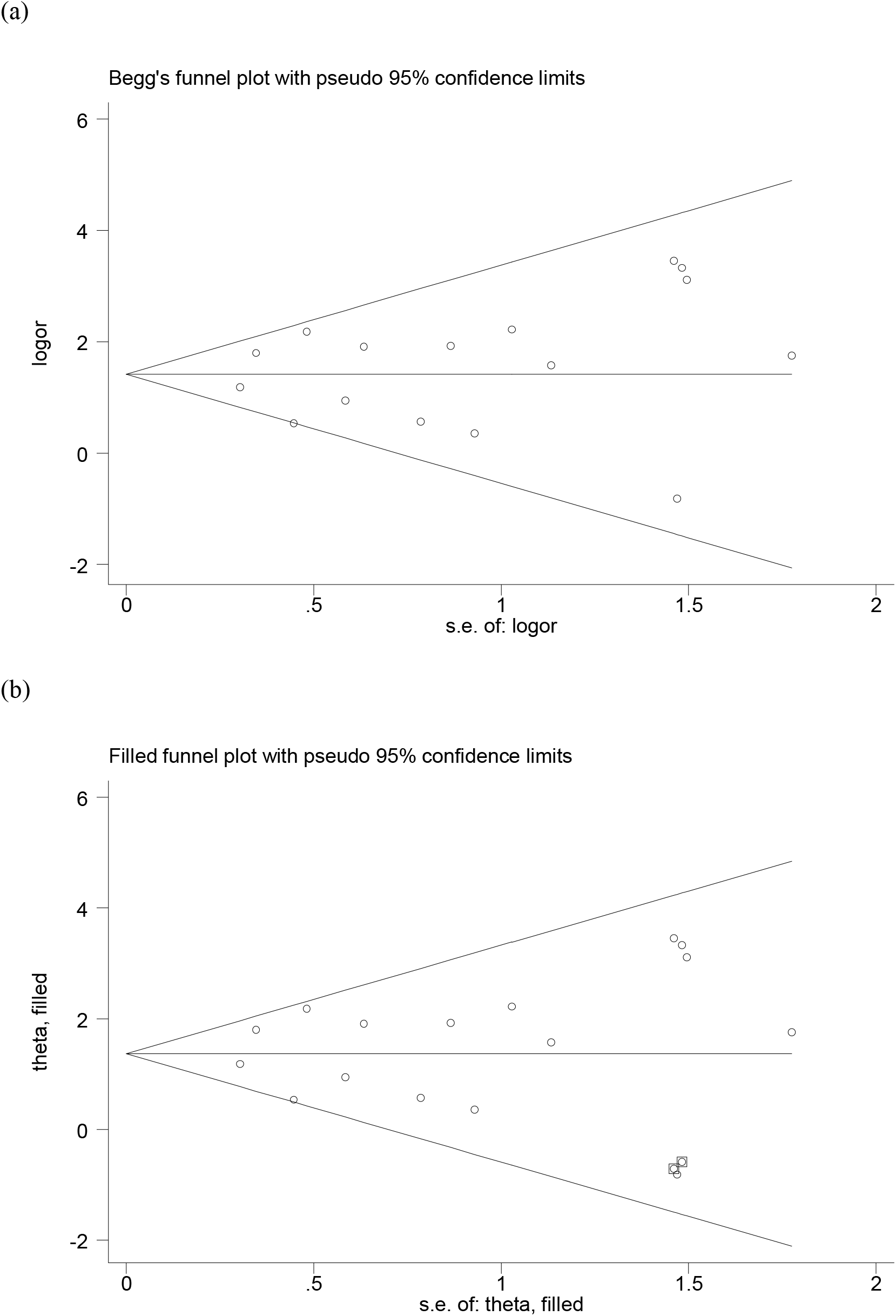
Funnel plot without (a) and with trim and fill (b) estimate for Cerebrovascular system diseases patients infected further with COVID-19 disease

**Supplementary Figure 6:**
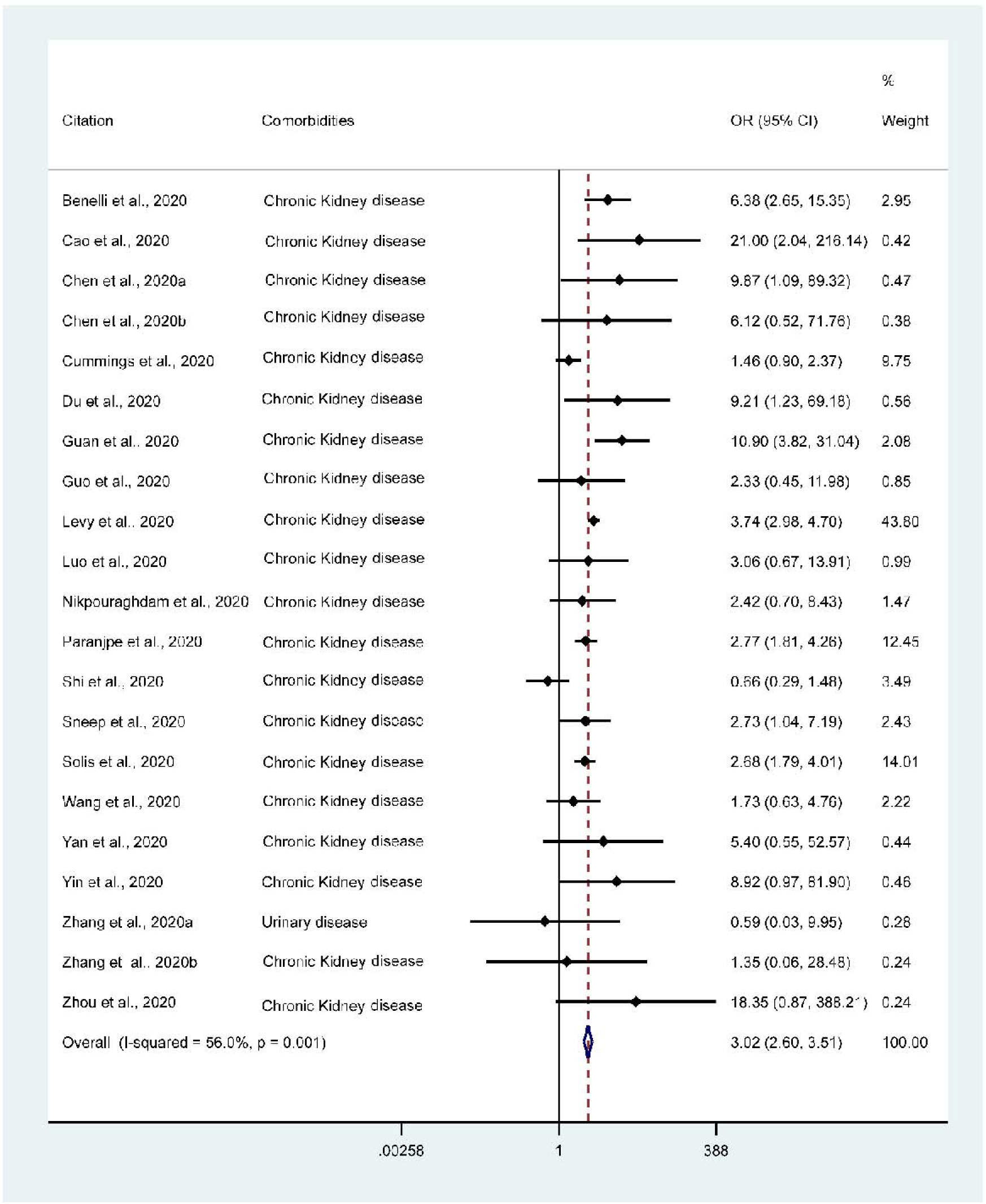
Likelihoods of death among patients with renal system diseases infected further with COVID-19 disease

**Supplementary Figure 6A:**
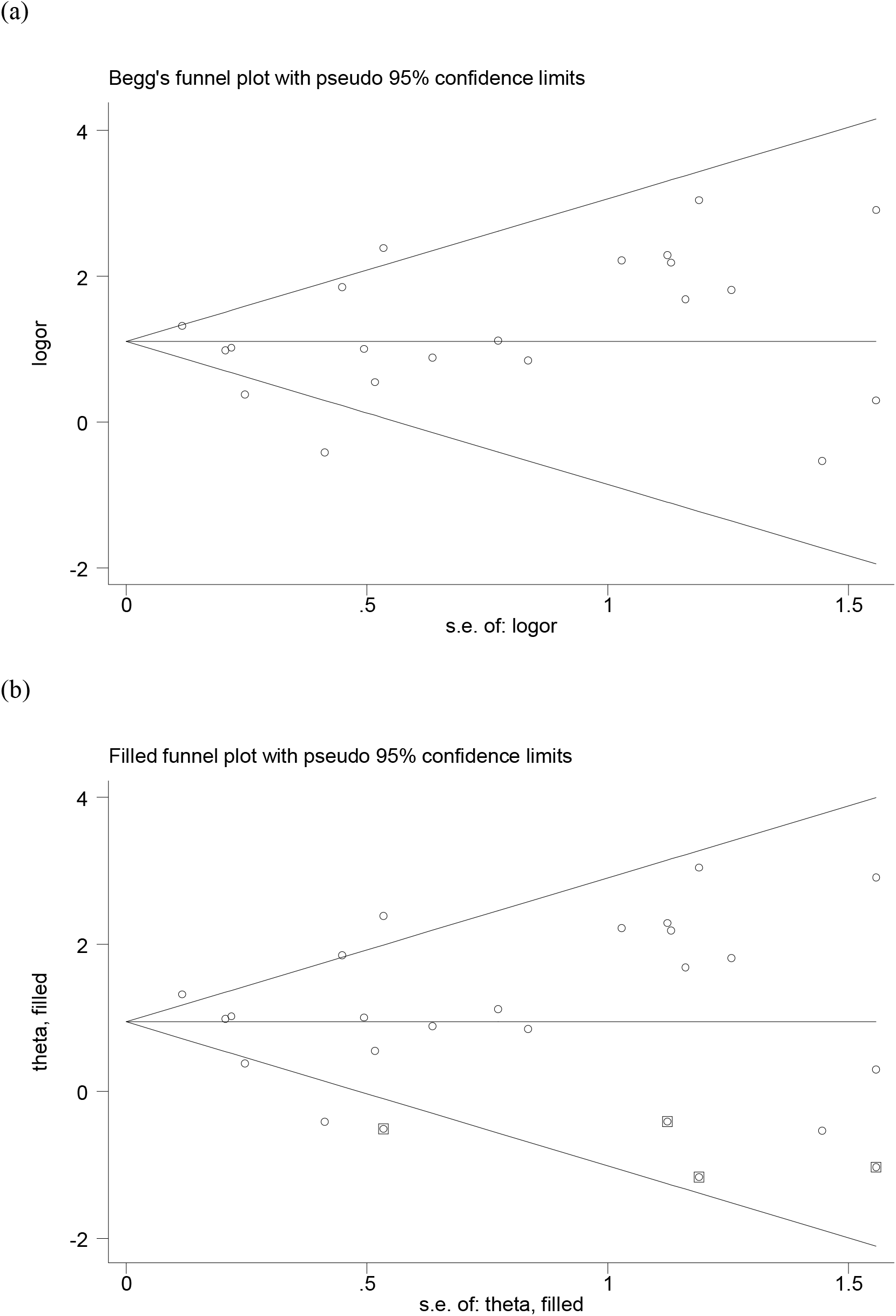
Funnel plot without (a) and with trim and fill (b) estimate for renal system diseases patients infected further with COVID-19 disease

**Supplementary Figure 7:**
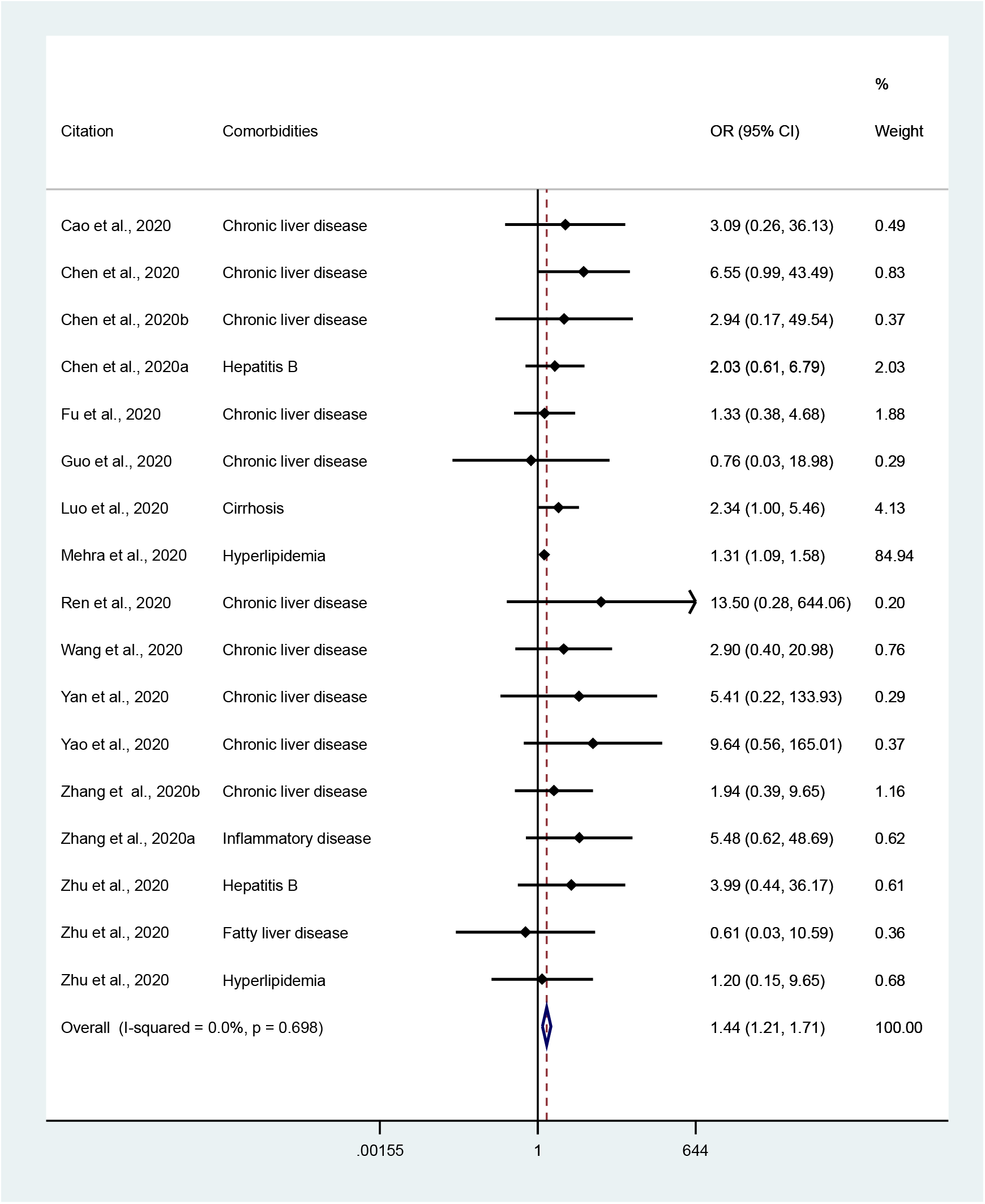
Likelihoods of death among patients with existing liver system diseases patients infected further with COVID-19 disease

**Supplementary Figure 8A:**
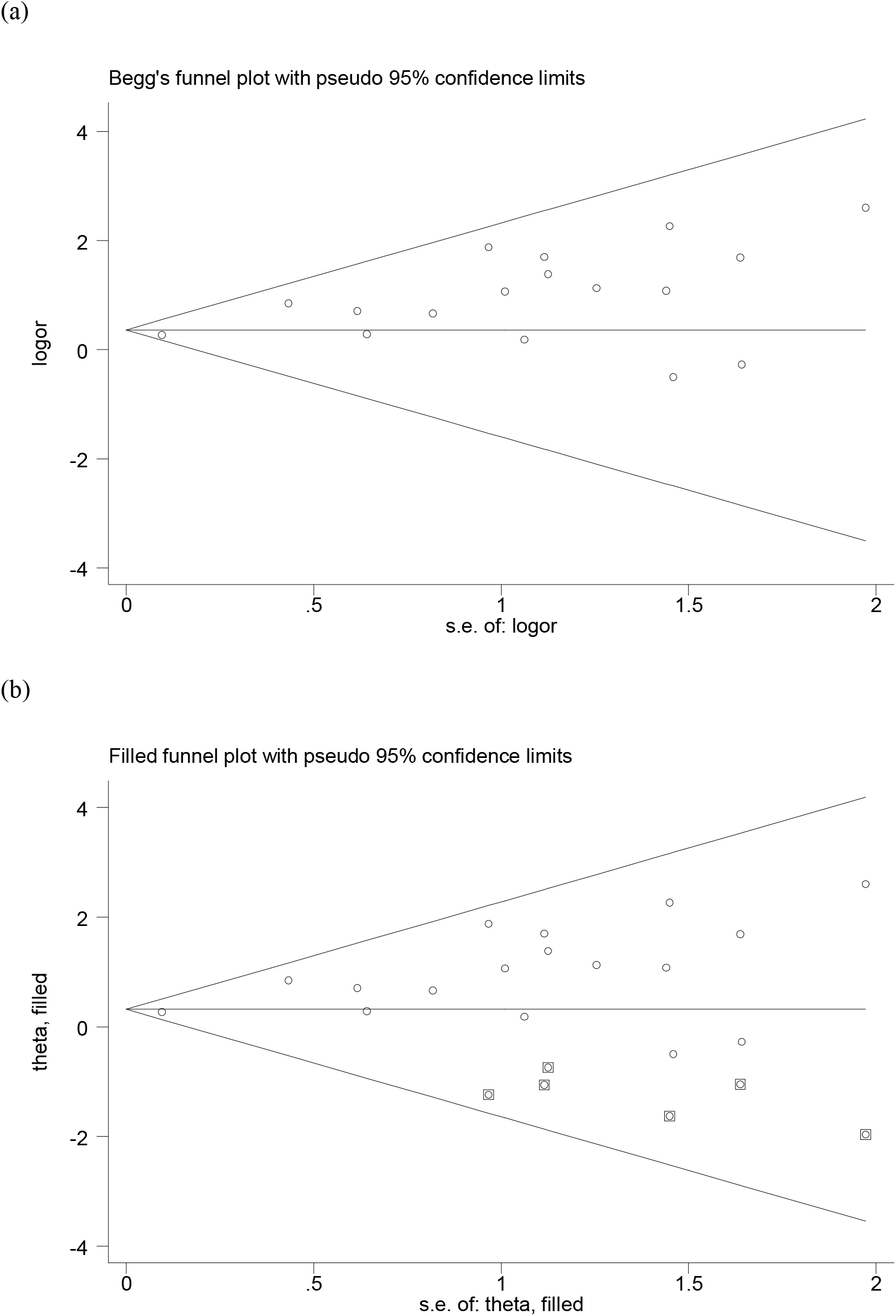
Funnel plot without (a) and with trim and fill (b) estimate for liver system diseases patients infected further with COVID-19 disease

**Supplementary Figure 8:**
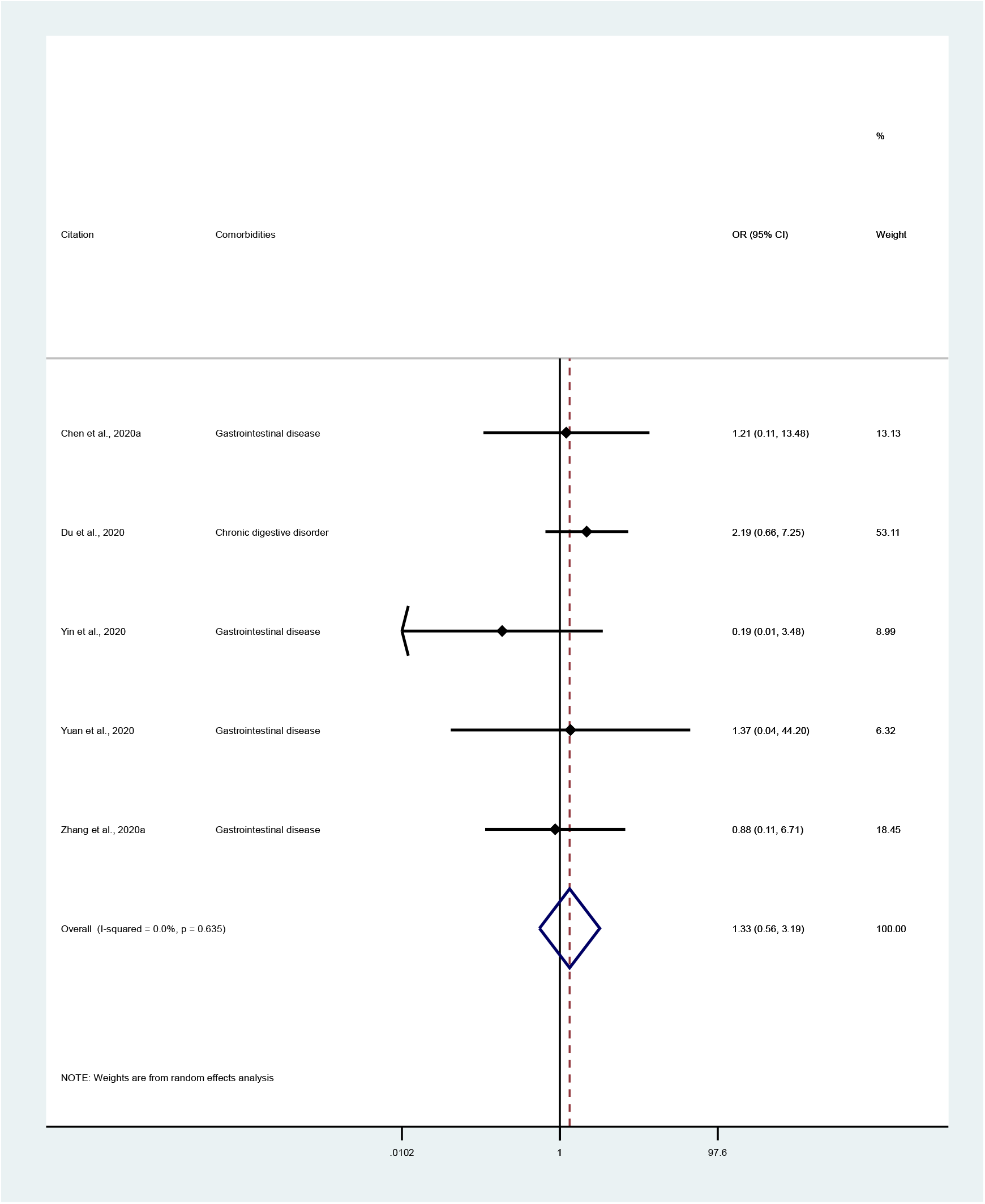
Likelihoods of death among patients with existing gastrointestinal system diseases patients infected further with COVID-19 disease

**Supplementary Figure 8a:**
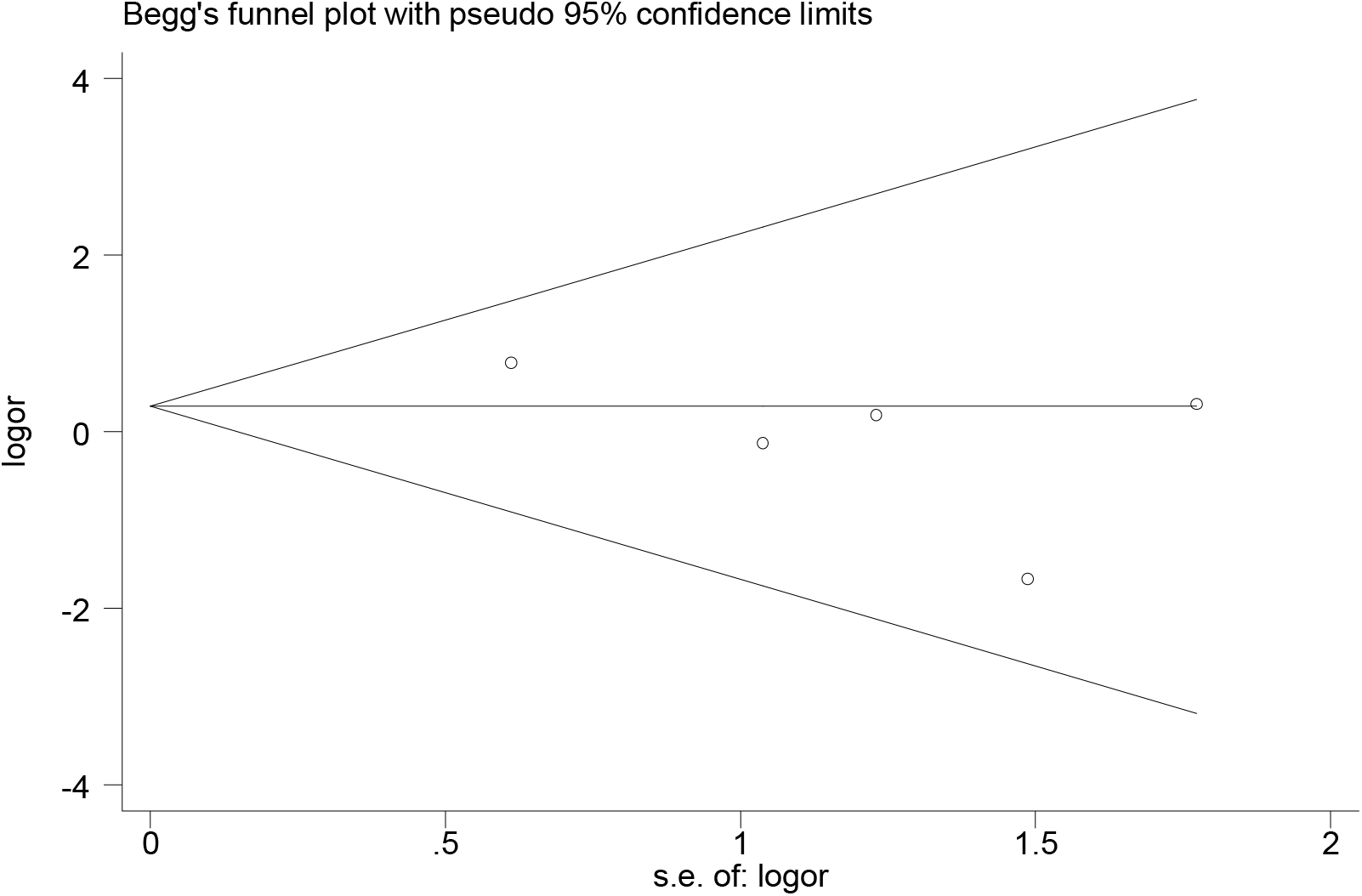
Funnel plot for gastrointestinal system diseases patients infected further with COVID-19 disease

**Supplemental Table 1.** Medline search results for pre-existing morbidities among COVID-19 patients

| # | Searches | Results |
| --- | --- | --- |
| 1 | (COVID-19 or 2019-nCoV or Coronavirus or SARS-CoV-2).mp. [mp=title, abstract, original title, name of substance word, subject heading word, floating sub-heading word, keyword heading word, organism supplementary concept word, protocol supplementary concept word, rare disease supplementary concept word, unique identifier, synonyms] | 17285 |
| 2 | (Comorbidit* or Morbidit*).mp. [mp=title, abstract, original title, name of substance word, subject heading word, floating sub-heading word, keyword heading word, organism supplementary concept word, protocol supplementary concept word, rare disease supplementary concept word, unique identifier, synonyms] | 573824 |
| 3 | (Mortalit* or Death or Died*).mp. [mp=title, abstract, original title, name of substance word, subject heading word, floating sub-heading word, keyword heading word, organism supplementary concept word, protocol supplementary concept word, rare disease supplementary concept word, unique identifier, synonyms] | 1826650 |
| 4 | 1 and 2 and 3 | 433 |
| 5 | limit 4 to (english language and yr="2019 -Current") | 146 |

**Supplemental Table 2.** CINAHL search results for pre-existing morbidities among COVID-19 patients

| # | Query | Limiters/Expanders | Last Run Via | Results |
| --- | --- | --- | --- | --- |
| S1 | “( covid-19 OR 2019-ncov OR coronavirus OR sars-cov-2 )<br>AND ( comorbidit* OR morbidit* ) AND ( mortalit* OR deat*<br>OR died ) | Limiters - Published Date: 20191201-20200431; English Language<br>Expanders - Apply equivalent subjects<br>Search modes - Boolean/Phrase | Interface - EBSCOhost<br>Research Databases<br>Search Screen -<br>Advanced Search<br>Database - CINAHL<br>Complete | 19 |

**Supplemental Table 3.** Web of science search results for pre-existing morbidities among COVID-19 patients

| Search terms | Results |
| --- | --- |
| (covid-19 OR 2019-ncov OR coronavirus OR sars-cov-2) AND TOPIC: (comorbidit* OR morbidit*) AND TOPIC: (mortalit* OR deat* OR died)<br>**Timespan:** 2019-2020. **Indexes:** SCI-EXPANDED, SSCI, A&HCI, CPCI-S, CPCI-SSH, ESCI, CCR-EXPANDED, IC | 64 |

**Supplemental Table 4.** SCOPUS search results for pre-existing comorbidities and the mortality risk in COVID-19

| Search strategy | Results |
| --- | --- |
| TITLE-ABS-KEY ( ( ( covid-19 OR 2019-ncov OR coronavirus OR sars-cov-2 ) AND ( comorbidit* OR morbidit* ) AND ( mortalit* OR death* OR died ) ) ) AND ( LIMIT-TO ( PUBYEAR , 2020 ) OR LIMIT-TO ( PUBYEAR , 2019 ) ) | 142 |

**Supplemental Table 5.** Narrative review for pre-existing morbidities and mortality risk among patients in COVID-19 infection.

| Study | Study design, Country | Sample | Results |
| --- | --- | --- | --- |
| Grasselli et al., 2020 | Retrospective cohort, Italy | Total of 1591 COVID-19 patients admitted into ICU in 72 hospitals from 20 February to 18 March 2020 | Around 49% of the patients had hypertension, 21% had cardiovascular disease, 18% had hypercholesterolemia, 17% had type-2 diabetes, and 8% had malignancy. A total of 405 (26%) patients died. Around 38% of the patients with hypertension were died in ICU as compared to 16% discharged. |
| Du et al., 2020 | Prospective cohort, China | Total of 109 died COVID-19 patients from three hospitals in Wuhan, China. Data were collected from 25 Dec to 15 February 2020 | COVID-19 patients who were died mostly had pre-existing hypertension (59.6%), cardiovascular disease (33.9%), diabetes (31.2%), digestive disorders (16%), chronic respiratory diseases (15.6%), malignancy (7.3%), chronic kidney disease (7.3%), 7.3% had peripheral vascular disease. |
| Zhang et al., 2020 | Retrospective cohort, China | Total of 82 died COVID-19 patients admitted Wuhan University's hospital from 11 January to 10 February 2020. | Patients died following secondary COVID-19 mostly had pre-existing hypertension (56.1%) following cardiovascular disease (20.7%), diabetes (18.3%), immunodeficiency (17.1%) chronic respiratory diseases (14.6%), cerebrovascular disease (12.2%), malignancy (7.3%), chronic kidney disease (4.9%), and chronic liver disease (2.4%). |
| Kim et al., 2020 | Retrospective cohort, Korea | Sample consisting of 101 deceased patients from February 19 to March 20, 2020 | COVID-19 patients who were died mostly had pre-existing hypertension (64.4%), cardiovascular disease (21.8%), diabetes (43.6%), digestive disorders (16%), chronic respiratory diseases (27.7%), dementia (25.7), dyslipidaemia (15.8%), cerebrovascular disease (15.8%), malignancy (15.8%), and renal diseases (14.8%). |
| Yao et al., 2020 | Retrospective cohort, China | Sample of 55 patients who died were collected from East Hospital of Wuhan University as of February 18, 2020 | Patients died following secondary COVID-19 mostly had pre-existing hypertension (60%) diabetes (26%), cardiovascular (31%), cerebrovascular disease (22%), malignancy (7%), chronic lung diseases (22%), chronic kidney disease (9%), and chronic liver disease (6%). |
| Cheng et al., 2020 | Prospective cohort, China | Total of 701 COVID-19 patients from China | Around 16.1% (113 person) died in hospital. Patients with acute kidney injury were reported higher risk of death with a gradual increase across stages of injury: stage 1 (HR, 3.51; 95% CI, 1.53-8.02), stage 2 (HR, 6.24; 95% CI, 2.73-14.27), stage 3 (HR, 9.81; 95% CI, 5.46-17.65). |

**Supplemental Table 6.** Newcastle-Ottawa scale assessment of study quality for **cross-sectional study**

| Author | Selection |  |  |  | Comparability |  | Outcome |  | Study quality |
| --- | --- | --- | --- | --- | --- | --- | --- | --- | --- |
|  | 1 | 2 | 3 | 4 | 5 |  | 6 | 7 |  |
|  | Representativeness of the sample | Sample size | Ascertainment of exposure | Non-respondents | The subjects in different outcome groups are comparable, based on the study design or analysis. Confounding factors are controlled. |  | Assessment of outcome | Statistical test is appropriate |  |
| Solis et al., 2020 | * | * | * |  | * |  |  | * | 5 |

**Supplemental Table 7.**
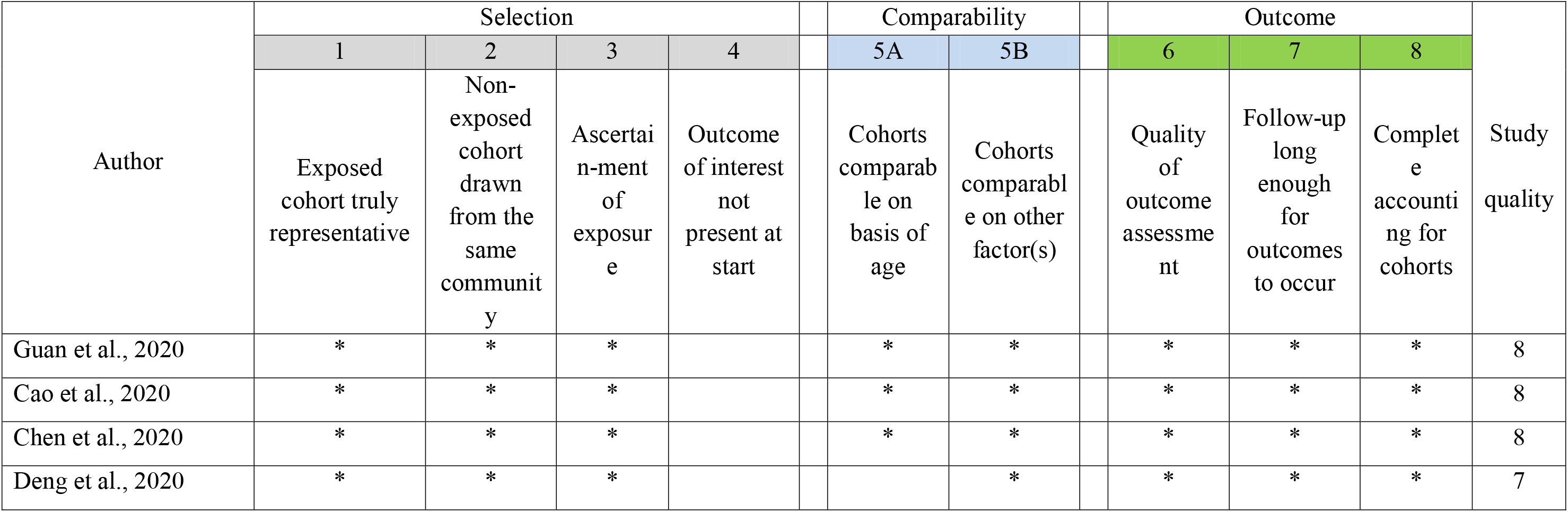
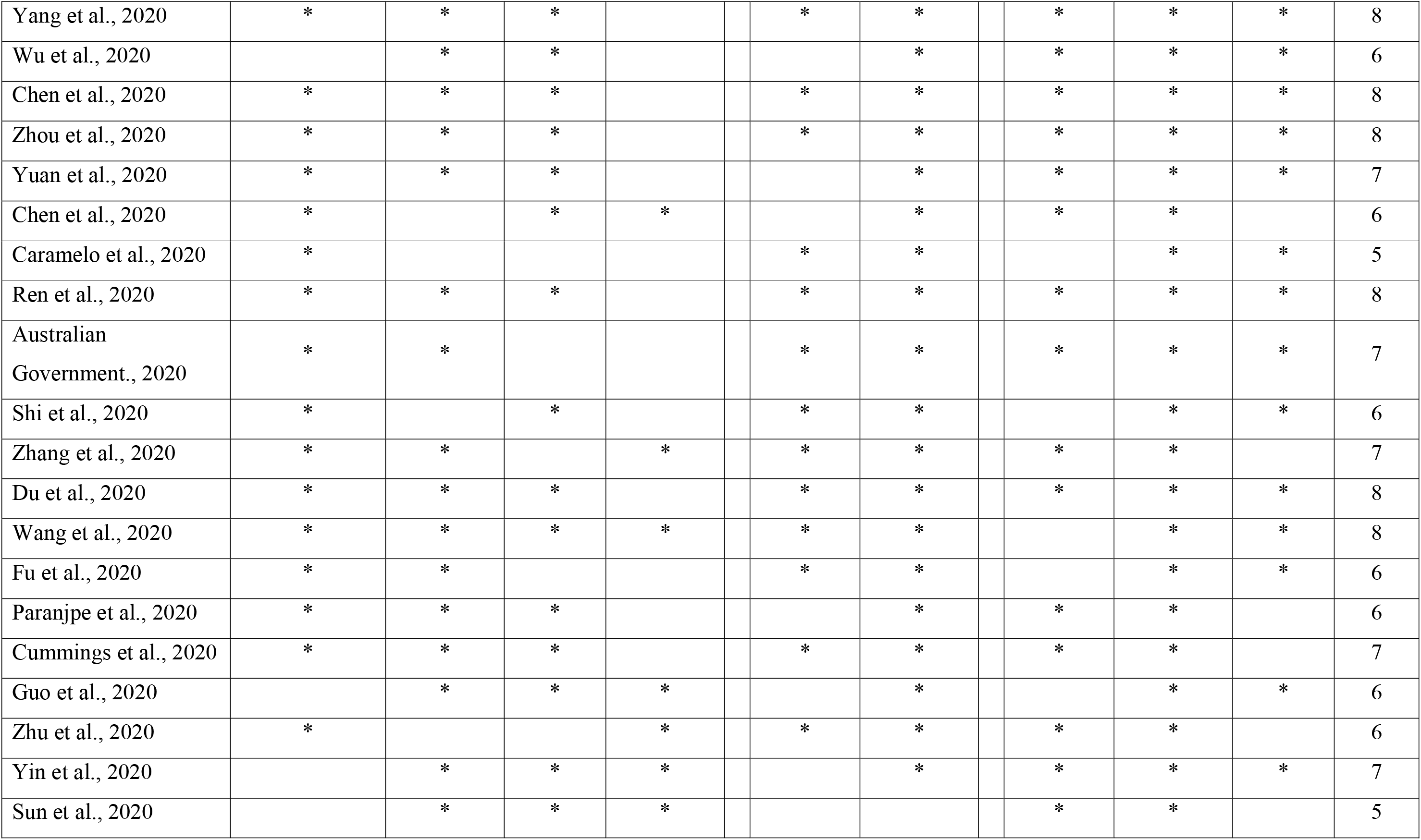
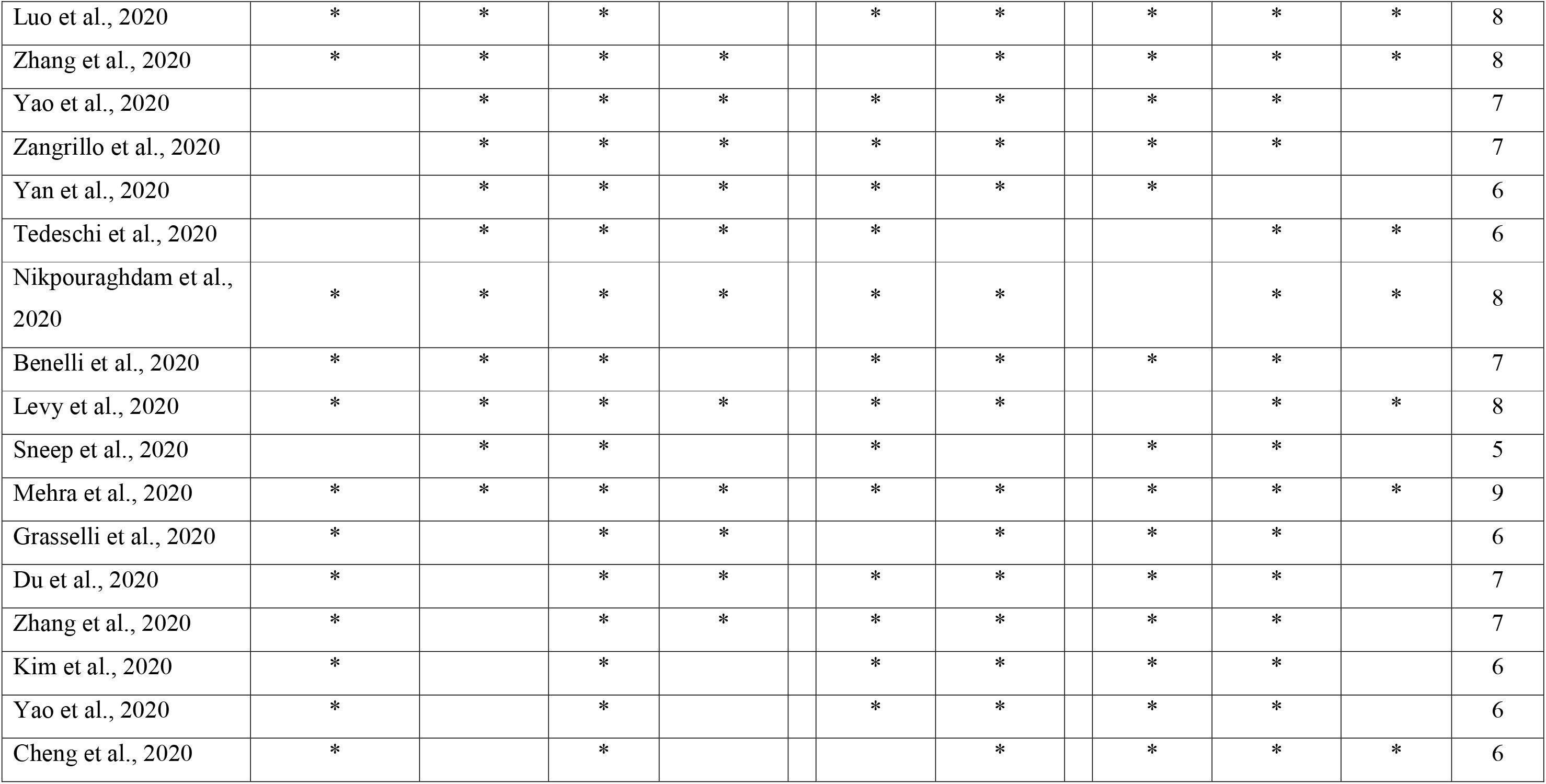
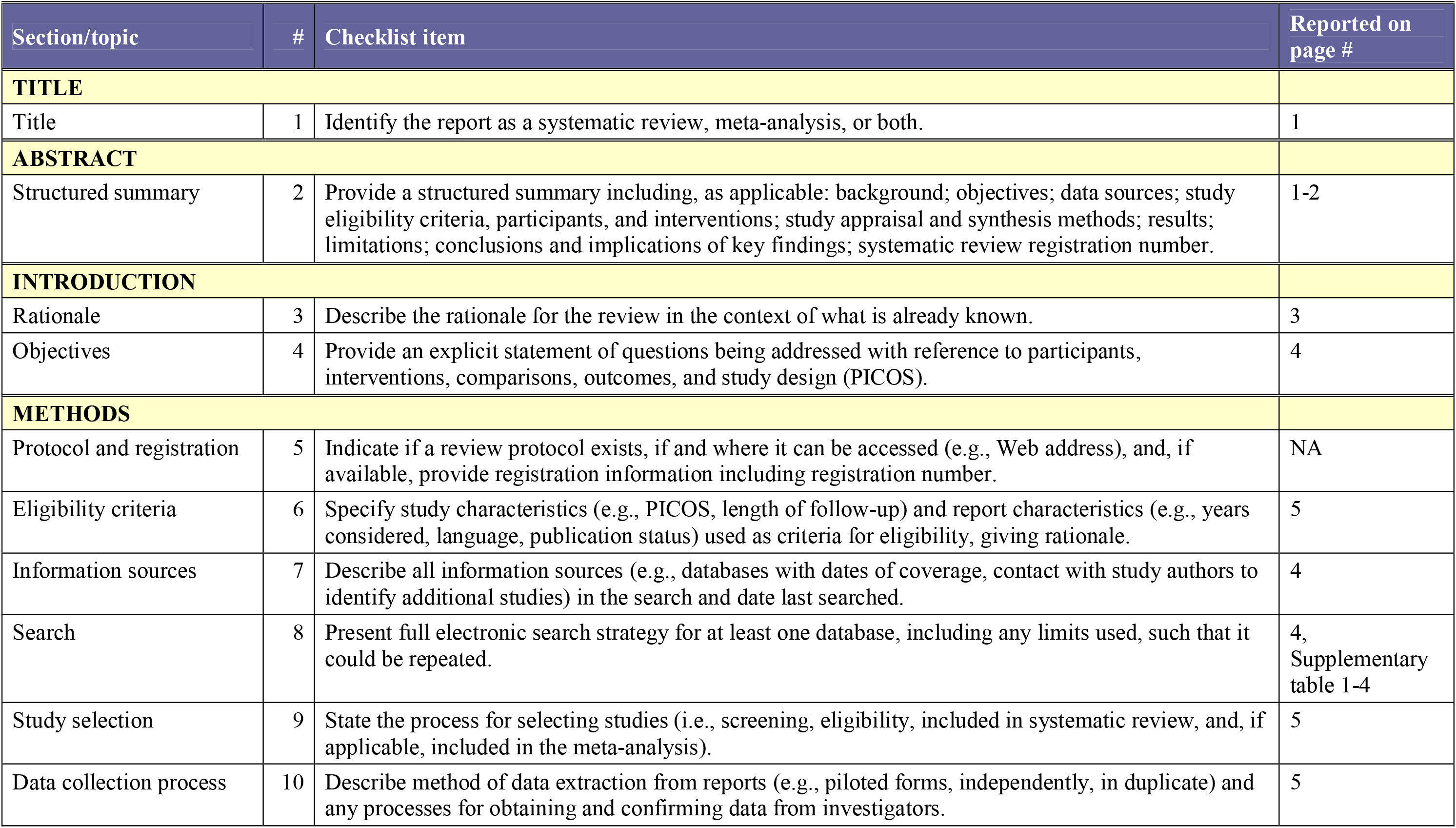
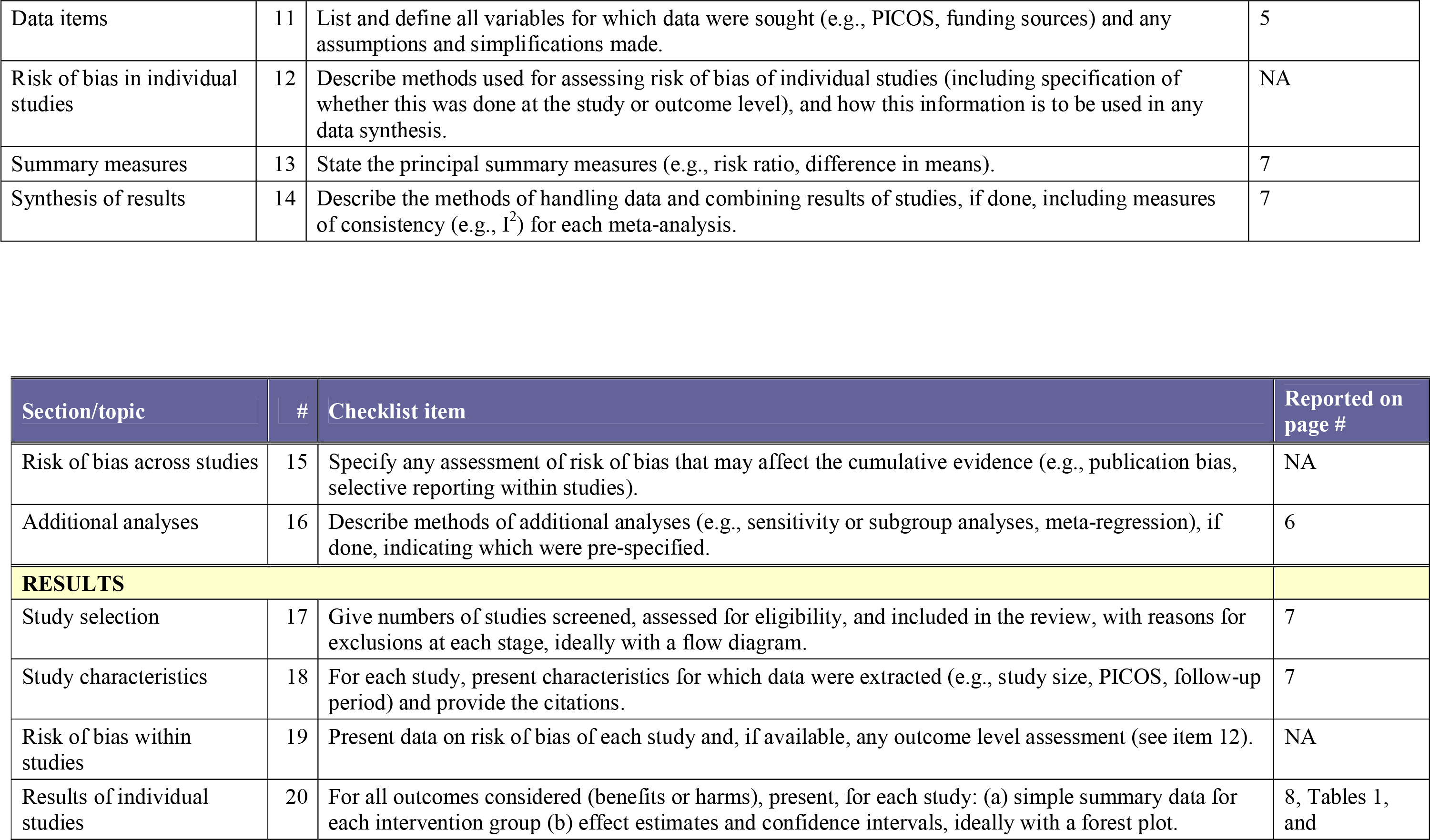
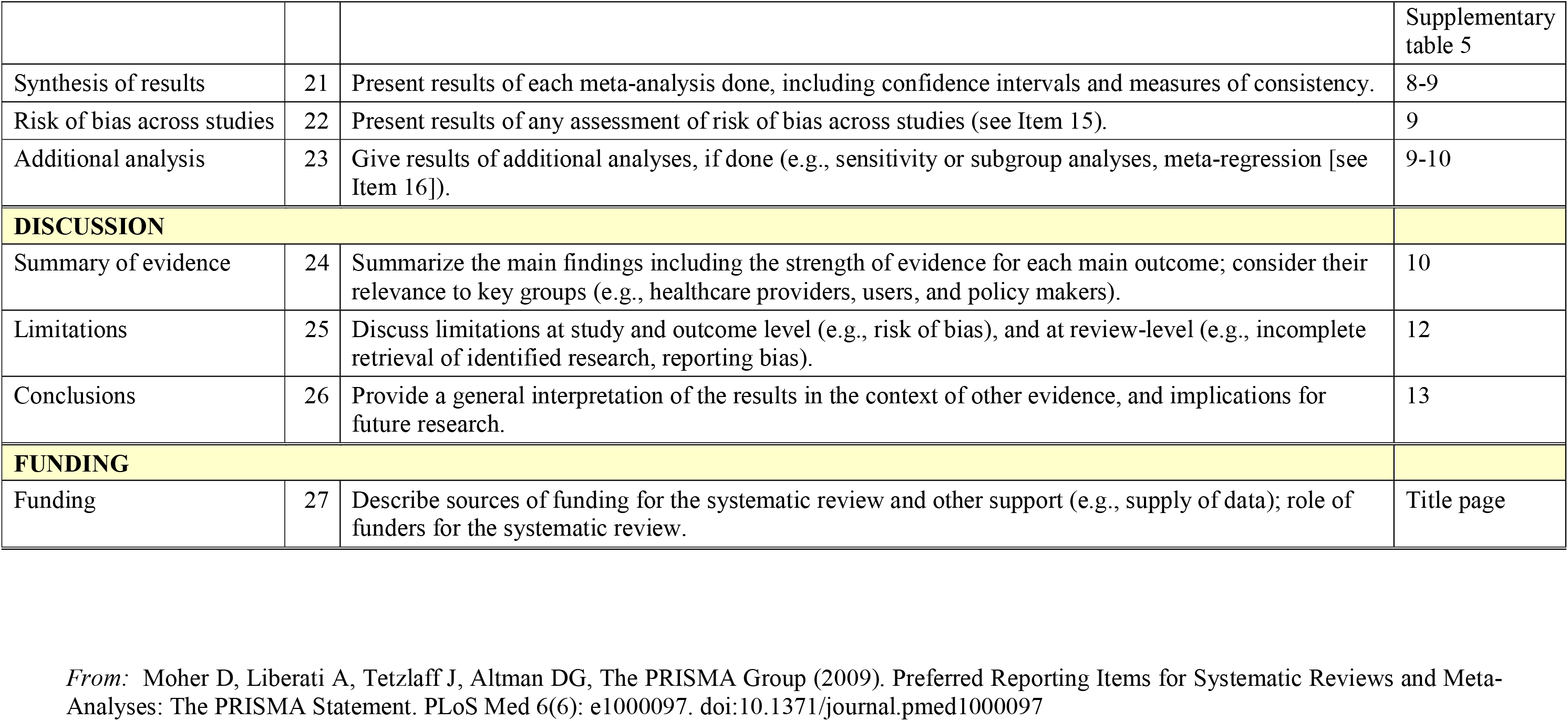
Newcastle-Ottawa scale assessment of study quality for **cohort study**

